# Multiscale Modelling Reveals Accelerating Community Outbreak Risks of Measles in the United States

**DOI:** 10.64898/2026.01.27.26345010

**Authors:** Siyu Chen, Ana I. Bento

**Affiliations:** Department of Public and Ecosystem Health, College of Veterinary Medicine, Cornell University

## Abstract

Measles resurgence in high-income countries that previously achieved elimination reveals a critical surveillance failure: current systems rely on county-level aggregates that obscure fine-scale clustering where outbreaks originate. We assembled the nationwide multiscale vaccination database spanning 45 US states (2013–2025), encompassing over 50,000 schools, 13,000 districts, and 3,000 counties. We developed a gravity-based transmission framework and demonstrate that school-level effective reproduction numbers crossed the epidemic threshold in 2022–2023, a transition invisible to aggregated surveillance. Average susceptibility doubled from approximately 5% to 10% following the COVID-19 pandemic. Three independent mechanisms drive aggregation bias: enrollment-weighted averaging systematically dilutes small under-vaccinated schools; gravity kernels attenuate transmission potential with coarser spatial scales; and clustering signal erasure converts heterogeneous risk landscapes into falsely uniform county averages. School district boundaries frequently misalign with counties, creating cross-boundary transmission corridors that push well-vaccinated counties above epidemic threshold through spillover alone. State trajectories diverge markedly, shaped by exemption policies and local infrastructure. Preventing measles re-endemicity requires surveillance systems operating at the spatial scale where vulnerability accumulates.

## Introduction

Measles, once considered on the path to global elimination, is resurging across a wide range of settings globally. In 2024, an estimated 11 million infections and 95,000 deaths occurred worldwide, with the vast majority of fatalities among children under five years of age in low-income counties with fragile health systems. Nearly 800,000 more cases than pre-pandemic levels [1]. Yet the resurgence is not confined to resource-limited settings. In high-income countries that had previously achieved elimination, are again experiencing sustained transmission and large outbreaks. In Italy, unvaccinated adults contribute substantially to measles transmission [2]. Canada lost elimination status in November 2025 after more than 12 months of continuous transmission [3]. In the United States, more than 2,000 confirmed cases and three deaths were reported in 2025, the highest annual total in over three decades [4].

These diverse settings share a common problem of inadequate vaccination coverage, but the epidemiology differs fundamentally between endemic and post-elimination contexts. In high-burden counties, high transmission intensity means unvaccinated children are infected early in life, with little time for the susceptible population to accumulate. In contrast, the absence of circulating virus post elimination allows under-vaccinated cohorts to age without exposure and natural infection, silently building a reservoir of susceptible individuals over years until an importation ignites an outbreak, most often among school-aged children [5-6].

As this reservoir grows, its spatial distribution becomes as consequential as its total number. Dispersed susceptible individuals can be buffered by immune neighbors, with local herd immunity interrupting transmission chains before they propagate. Whereas spatially clustered pockets of under-vaccination can drive explosive local transmission even when regional averages coverage remains near the 95% threshold commonly associated with herd immunity.

Current surveillance systems in the United States are poorly equipped to distinguish these contrasting scenarios. Routine monitoring of measles-mumps-rubella (MMR) coverage replies predominantly on state- and county-level aggregates complied from the National Immunization Survey and school entry assessments [7-9]. These metrics provide valuable population-level estimates but obscure fine-scale heterogeneity at the level where clustering of susceptible occurs in individual schools and their surrounding communities, where outbreaks are seeded and sustained.

Theoretical studies have long suggested that spatial clustering of non-vaccination substantially increases local outbreak risk and that aggregation to coarse spatial units can systematically underestimate this risk [10-11]. However, most prior work has relied on stylized simulations with idealized spatial structures, and empirical validation using actual school-level vaccination data across diverse geographic and policy contexts has been lacking.

In post-elimination settings such as the contemporary United States, susceptibility is concentrated among school-age children who were never exposed to circulating virus and who missed one or more doses of MMR. These children spend most of their day at school during term time, which functions as discrete mixing environments characterized by intense, repeated contact among peers. They typically remain in the same community throughout their primary and secondary education, with fewer than 4% of school-age children moving across county lines each year [12]. From a transmission perspective, schools are therefore the natural units of susceptibility clustering for evaluating measles transmission potential. Whereas school districts, which link multiple schools through shared administration, transportation, and catchment areas, represent functionally connected populations across which disease can readily propagate [13]. Importantly, school district boundaries often do not align with county boundaries. A single district may span multiple counties, and a single county may encompass portions of many districts, creating spillover pathways between counties that are invisible to county-level surveillance. Despite the central role of schools and districts in shaping measles transmission risks, multiscale mapping of susceptibility at these resolutions has been impeded by the decentralized and heterogeneous nature of US immunization data reporting, with each state maintaining its own system of varying completeness and accessibility (Table S1).

The spatial dynamics of measles transmission have been most extensively studied during the transition toward elimination, when high vaccination coverage created a patchwork of largely immune populations punctuated by small pockets of susceptible population [14-15]. In that regime, spatial coupling, whether hierarchical from large cities or lateral among neighboring towns, governs where infection persists and how it spreads following local fade-out [16]. Pre-elimination and pre-endemicity settings may exhibit a mirror-image structure: the former characterized by transmission fading out spatially, the latter by vulnerability building up silently. The central question shifts from where infection persists to where susceptibility accumulates. The underlying spatial architecture may be similar, but the observable signal is not. Understanding this transition from elimination back toward endemicity carries profound implications for public health policy. In endemic or transitional regimes, spatial coupling can be inferred from correlated epidemic trajectories across locations [14], where in post-elimination settings transmission is largely absent and susceptible populations accumulate silently. Whether this accumulation is spatially synchronous—clustered vulnerability rising in concert across connected communities—or asynchronous, with pockets of high risk buffered by immune neighbors, remains unknown. Gravity models have proven powerful for capturing how spatial connectivity structures transmission dynamics in the presence of circulating pathogens [17] but have not been adapted to quantify how the same connectivity shapes the build-up of transmission risk in the absence of ongoing infection.

Here, we address this gap by assembling a nationwide, multiscale database of MMR vaccination coverage spanning 45 US states from 2013 to 2025, integrating more than 50,000 schools, 13,000 districts, and 3,000 counties. This resource enables the first empirical multiscale mapping of vaccination coverage and transmission risks in the United States and allows direct quantification of aggregation bias using real-world data across diverse geographic and policy environments. Building on this dataset, we develop a multiscale modelling framework incorporating gravity-based mixing kernels, age-place contact structures, and cross-boundary school-district spillovers to quantify risk synchrony across scales and identify threshold-crossing dynamics invisible to conventional surveillance. We find susceptibility has nearly doubled post-pandemic, with divergent state trajectories, exposing hidden hotspots of transmission potential that current systems fail to detect. These findings reveal that the spatial architecture of US measles vulnerability has fundamentally shifted, with profound implications for outbreak preparedness amid ongoing global resurgence.

## Results

### Multiscale vaccination coverage reveals hidden vulnerability

Measles-Mumps-Rubella (MMR) vaccination coverage of kindergarteners exhibited broad geographic heterogeneity across three nested spatial scales: county, school district, and individual school in the United States for the 2024-2025 school year (or states for the 2023–2024, Figure S1 for observations by state and year) (Figure 1 A, C, E and G). At the county level, coverage ranged from 70% to over 98%, with increasingly fine-grained pockets of low coverage (<80%) emerging at the district and especially school levels (Figure 1 A, C, E and G).

**Figure 1.**
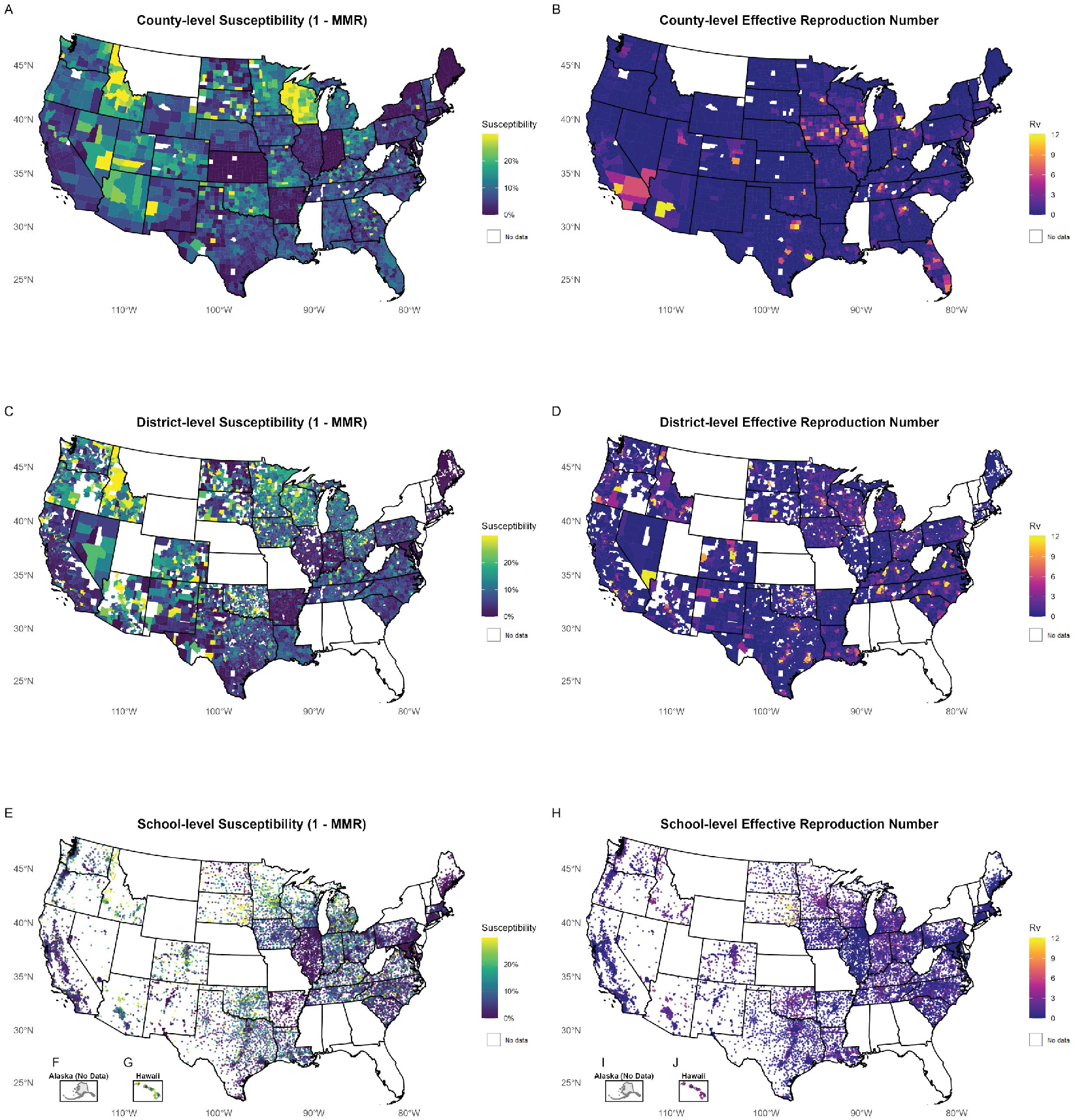
Multiscale spatial distribution of Measles-Mumps-Rubella (MMR) vaccination coverage and transmission potential in United States. (A, C, E and G) shows MMR coverage at the county level, school district level and individual school level for the 2024-2025 school year (or for the 2023-2024 school year in 12 states) respectively. Coverage is displayed as the proportion of kindergarten students with documented MMR vaccination. (B, D, H and J) show estimates of effective reproduction numbers (*R*_*v*_) calculated using a gravity kernel transmission model, calibrated to R_0_ = 15 for measles, at the county level, school district level and individual school level for the 2024-2025 school year (or for the 2023-2024 school year in CA, ID, MD, OK, PA, SD states). F-G and I-J shows Alaska and Hawaii MMR coverage and *R*_*v*_ estimates. White areas indicate states without publicly available data at the respective spatial scale.

Despite deriving from identical underlying data, a higher proportion of counties fell below the 95% coverage threshold than individual schools in many states (Table S1). This arises because large schools have higher coverage than small, and aggregation dilutes the contribution of small, under-vaccinated schools (Figure S2). These patterns demonstrate that spatial aggregation systematically obscures coverage heterogeneity, and that apparent differences across scales are confounded by population size distributions.

When mapped to vaccination-adjusted effective reproduction numbers, *R*_*v*_, quantifying transmission potential using a gravity-based mixing kernel model calibrated to a measles basic reproduction number, *R*_*0*_ of 15 (Figure 1 B, D, H and J), risk appears sparse at the county scale, but numerous, spatially clustered pockets exceeding the epidemic threshold (*R*_*v*_ >1) emerge at the district and school scales. Several states with high county-average coverage still harbor many districts and schools above the epidemic threshold, underscoring the potential for sustained transmission despite reassuring county averages.

Overall, the contrast between scales shows clear aggregation bias: county-level summaries dilute localized susceptibility clusters that drive transmission potential. The progressive disaggregation from county to school level exposed critical transmission risk clusters that were not detectable at coarser spatial resolutions, highlighting the importance of fine-scale surveillance for identifying communities vulnerable to measles outbreaks.

### Susceptibility–*R*_*v*_relationships attenuate with aggregation

In Figure 2A, we examined the relationship between susceptibility (100% - MMR coverage) and disease transmission potential measured by *R*_*v*_ across three spatial scales: school, school district, and county levels in 31 states for the 2024-2025 school year (or for 2023-2024 according to data availability). There are three essentially independent effects that emerge (Figure 2A). First, susceptibility-*R*_*v*_ relationship weakens with aggregation. Higher susceptibility is consistently associated with higher transmission potential, as depicted by the increasing black regression lines in all three scales. The regression line is steepest at school level and shallowest at county level although the red dashed vertical lines, indicating average susceptibility, sit around 7-8% across all three scales; this indicates the susceptibility-*R*_*v*_ relationship weakens as coverage is aggregated to coarser spatial units. Nevertheless, average coverage (∼92-93%) remains stable regardless of aggregation level.

**Figure 2.**
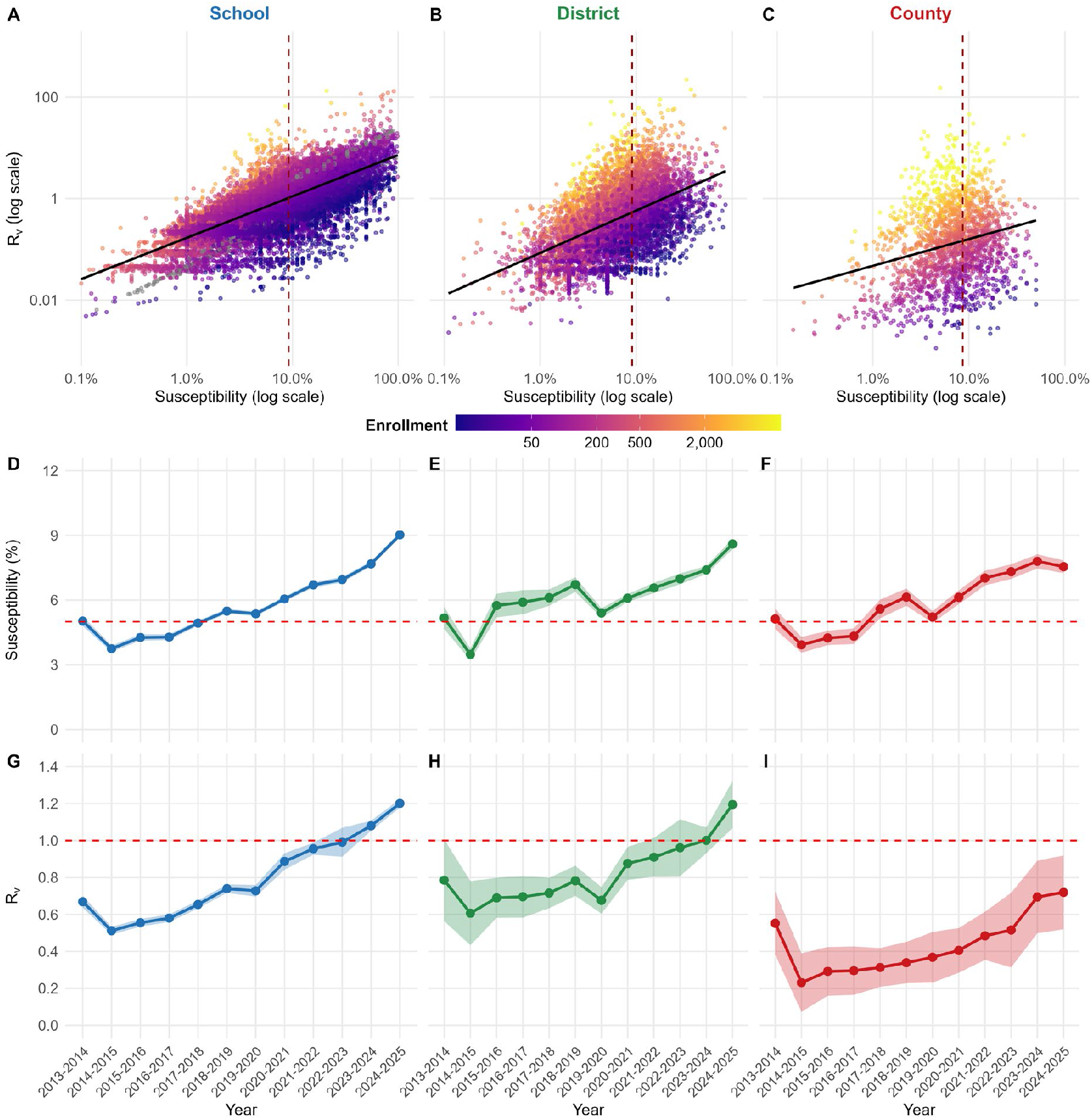
Scale-dependent susceptibility and temporal trends in transmission potential. (A-C) Relationship between susceptibility (100%−MMR coverage) and the estimates of effective reproduction number (*R*_*v*_) at the individual school level, school district level, and county level in the log–log axes among 31 states that report MMR coverage data at all three levels for the 2024-2025 school year (AR, AZ, CO, CT, IA, IL, IN, KY, LA, MA, ME, MI, MN, NC, ND, NM, NV, OH, OR, SC, TN, TX, VA, WA, WI or 2023-2024 school year for CA, ID, MD, OK, PA, SD). Each point represents an individual unit, colored by enrollment. Solid black lines represent a simple linear regression fit between *R*_*v*_ and susceptibility at the log scales. The vertical dashed line marks the mean susceptibility for each scale while grey dashed curves represent mean *R*_*v*_ at a certain level of susceptibility. (D–F) Time course of average susceptibility (points and solid lines) with 95% confidence (shaded ribbons, calibrated with mean ±1.96×standard deviation) at the individual school, school district, and county level from 2013-2014 to 2024-2025 school year among states. The red horizontal dashed line denotes the susceptible pool threshold, susceptibility = 5%. (G-I) Time course of average *R*_*v*_ (points and solid lines) with 95% confidence (shaded ribbons, calibrated with mean ±1.96×standard deviation) at the individual school, school district, and county level from 2013-2014 to 2024-2025 school year among states. The red horizontal dashed line denotes the epidemic threshold, Rv = 1.0.

Second, distance-driven attenuation of *R*_*v*_ across scales. *R*_*v*_ decreases from the scale of schools to that of districts and even more so, to the county scale, as visible in the downward shift of the entire cloud of points from one scale to the other. This is directly attributable to the greater distance between districts, compared to the distance among schools in a single district, and the even greater distances between counties, because the gravity kernel of the *R*_*v*_ function weights transmission proportional to exp(−distance).

Third, enrollment-driven changes in *R*_*v*_ distribution. The role of the small schools, with smaller *R*_*v*_, at the level of individual schools, is greatly attenuated at the district level, as these schools are amalgamated into districts containing several schools, so that total enrollment increases accordingly. This tendency is continued at the county level where the small schools almost “disappear”, and the overall enrollment becomes much larger. This process is reflected in changes, from school level to district level to county level, of the color gradient of dots from bottom to top in each panel. The color gradient – blue-green–yellow– reflects that *R*_*v*_ scales up in proportion to enrollment as a direct consequence of the gravity kernel; thus it might be expected that the enrollment effect would produce an overall net increase in *R*_*v*_, but the effect of greater inter-county distances clearly predominates in producing the downward shift of the cloud of points in the county level display.

Temporal trends revealed a marked increase in population susceptibility (Figure 2D-F and Figure S3A-C) and transmission potential (Figure 2G-I and Figure S3D-F) at all three scales following the COVID-19 pandemic. Average susceptibility remained relatively stable at approximately 4%– 5% from 2014 to 2019, near the critical 5% threshold corresponding to 95% herd immunity coverage. However, beginning in 2019–2020 and accelerating through the COVID-19 pandemic period, average susceptibility increased markedly, reaching approximately 8%–10% by 2024– 2025, nearly double the pre-pandemic levels and well above the herd immunity threshold.

### School-level *R*_*v*_crosses epidemic threshold

This decline in vaccination coverage translates directly into elevated transmission potential. At the school level, average *R*_*v*_ rose steadily from approximately 0.6 in 2013–2014 to exceed 1.0 by 2022– 2023, indicating that average transmission conditions now support sustained measles outbreaks (Figure 2G). Average school-level *R*_*v*_ continued to rise through 2024–2025, reaching approximately 1.2. District-level average *R*_*v*_ followed a parallel but attenuated trajectory, increasing from approximately 0.5 to 0.9 over the study period, approaching but not yet crossing the epidemic threshold (Figure 2H). County-level average *R*_*v*_ showed greater year-to-year variability but remained below 1.0 throughout (Figure 2I).

This scale-dependent attenuation indicates that school-level *R*_*v*_ captures early signals of spatial transmission that are progressively diluted by aggregation. The divergence between school-level *R*_*v*_ (now above threshold) and county-level *R*_*v*_ (still below threshold) underscores the limitation of coarse surveillance and the importance of fine-scale surveillance: monitoring at the county level alone would fail to detect that average conditions in schools have already crossed into a regime supporting sustained transmission.

### Clustering of susceptibility and scale-dependent risk

To assess how spatiotemporal clustering and dispersion of susceptible population influence transmission potential, we analyzed the relationship between within-state spatial autocorrelation of susceptibility measured by Geary’s C and average *R*_*v*_ across state-year combinations at each spatial scale (Figure 3A-C). At the school level, most state–years showed clear but moderate clustering (0.8< Geary’s C <1.0), while *R*_*v*_ values were fairly uniformly distributed between 0.5< *R*_*v*_ <2.5. A significant negative correlation was observed (*r* = −0.45, *p* < 0.001), indicating that state–years with more dispersed susceptibility patterns (Geary’s C > 1) had lower transmission potential.

**Figure 3.**
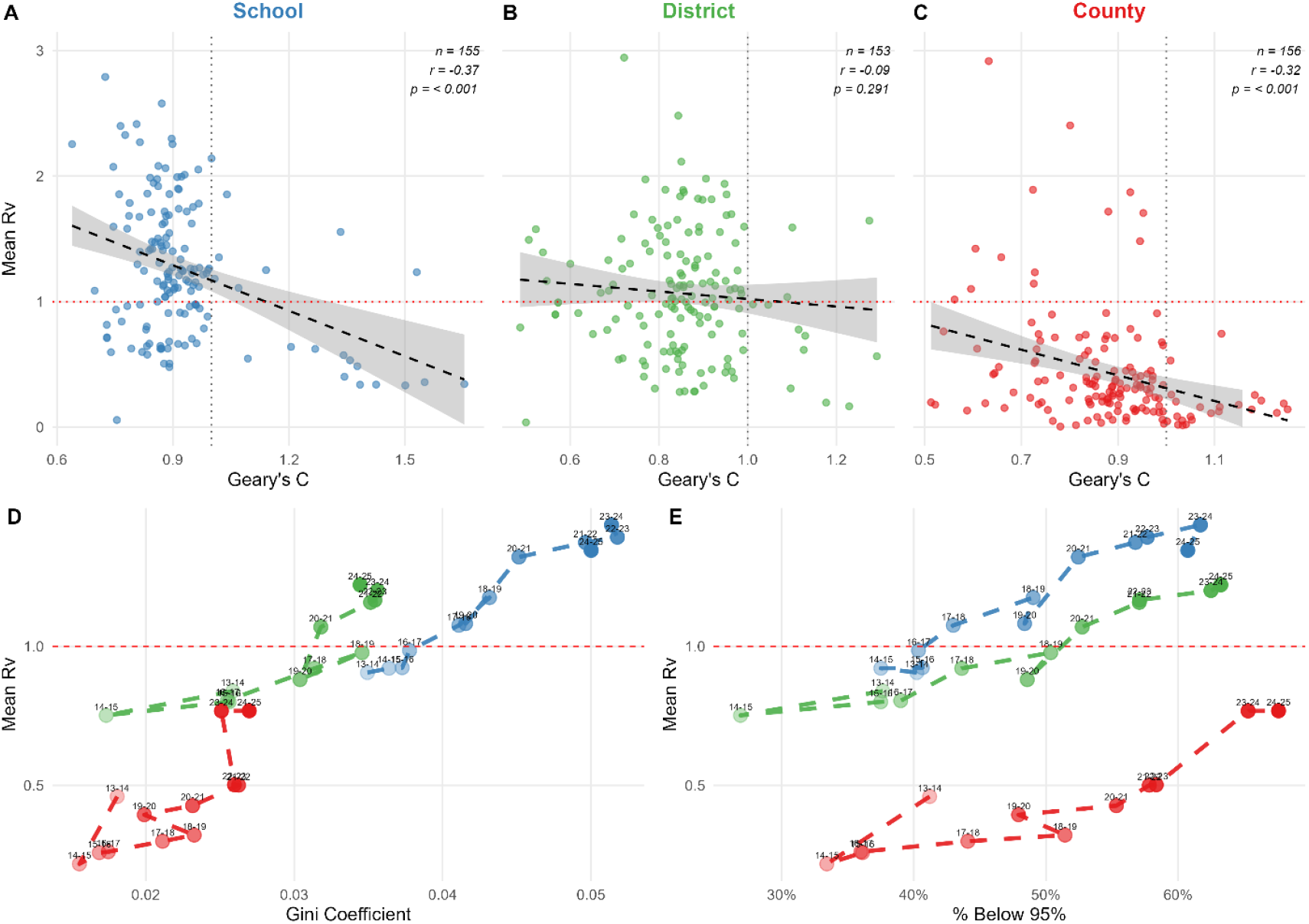
Spatiotemporal clustering of susceptibilities, vaccination coverage inequality, and their associations with transmission potential across scales. (A-C) Relationship over time between spatial clustering of susceptibility measured by Geary’s C and disease transmission potential measured by mean *R*_*v*_ within states, calculated separately at individual school (red), school district (blue), and county (green) levels respectively. Each point represents one state-year combination. Geary’s C < 1 represents under-vaccinated schools or school districts or counties clustering together, creating concentrated pockets of susceptibility; Geary’s C > 1 represents under-vaccinated schools, or school districts or counties dispersed or isolated among well-vaccinated neighbors, suggesting susceptibility is diluted spatially; Geary’s C = 1 represents no spatial autocorrelation. Number of state-year (*n*), Pearson correlation coefficients (*r*) and p-values are shown for each scale. *p* < 0.05 represents Pearson correlation coefficient significantly different from zero. Dashed black lines and shaded area show linear regression fits with 95% confidence bands; horizontal dotted red lines mark *R*_*v*_ = 1; vertical dotted black lines mark Geary’s C = 1. Temporal trajectories of mean *R*_*v*_ against vaccination coverage inequality measured by Gini coefficient (D) and proportion of schools or school districts or counties below the 95% herd immunity trajectory given R_0_=15 for measles (E). Red for county, green for school district and blue for school. Line segments connect successive years with labels indicating school years. Larger Gini coefficient represents greater disparity in vaccination coverage among schools or among school districts or among counties. Note that Gini coefficients are calculated separately within each spatial scale and are not directly comparable across scales; aggregation from schools to counties compresses the coverage distribution, systematically reducing observable inequality. Trajectories should be compared across years within each scale rather than across scales at the same Gini value.

Clustering strengthened in many state-year combinations distributed in the 0.5< Geary’s C <1 at the district and county levels, but aggregation reduced *R*_*v*_ values overall. The Geary’s C–Rv relationship attenuated at coarser scales: at the district-level the association was weak and non-significant (*r* = −0.11, *p* = 0.215), whereas at the county-level a moderate correlation persisted (*r* = −0.32, *p* <0.001). These scale-dependent patterns suggest that school-level clustering reveals elevated transmission potential that is partly obscured data are aggregated to districts and counties. At the county level, *R*_*v*_ is concentrated below 1 despite spatial autocorrelation, meaning that aggregation of school-level data masks the fact that many individual schools have *R*_*v*_ >1.

Inequality in vaccination coverage grew progressively indicated by increasing Gini coefficient accompanied by higher average *R*_*v*_ (Figure 3D). Notably, school-level trajectories (blue) consistently occupied higher *R*_*v*_ values than district (green) or county (red) trajectories at comparable levels of excess over herd immunity threshold (Figure 3E), reinforcing how fine-scale analysis detects elevated transmission risk that remains invisible at aggregated scales.

### Multidimensional scaling reveals states positions shift across scales, but clusters persist

Most of the variation of states in the matrix of correlation among five variables (Figure S4-S5), average vaccination coverage, median vaccination coverage, *R*_*v*_, Geary’s C, and Gini coefficient, was captured by the multidimensional scaling on only two dimensions, explaining >94% variance, at the school, school district and county levels (Figure 4). The first dimension basically summarizes the two closely correlated variables, average coverage and *R*_*v*_ (Figure 4A-C; Figure S6). The dots representing the states are colored according to *R*_*v*_ in the top three panels, showing a clear order from left to right (Figure 4A-C). The second dimension reflects the spatial autocorrelation of susceptibility, Geary’s C and well as *R*_*v*_. The dots representing the states are colored according to Geary’s C (Figure 4D-F), showing a clear order from top to bottom. The impression from Figure 1, that the states differ not only by overall coverage but by different internal patterns, is statistically confirmed by the multidimensional scaling.

**Figure 4.**
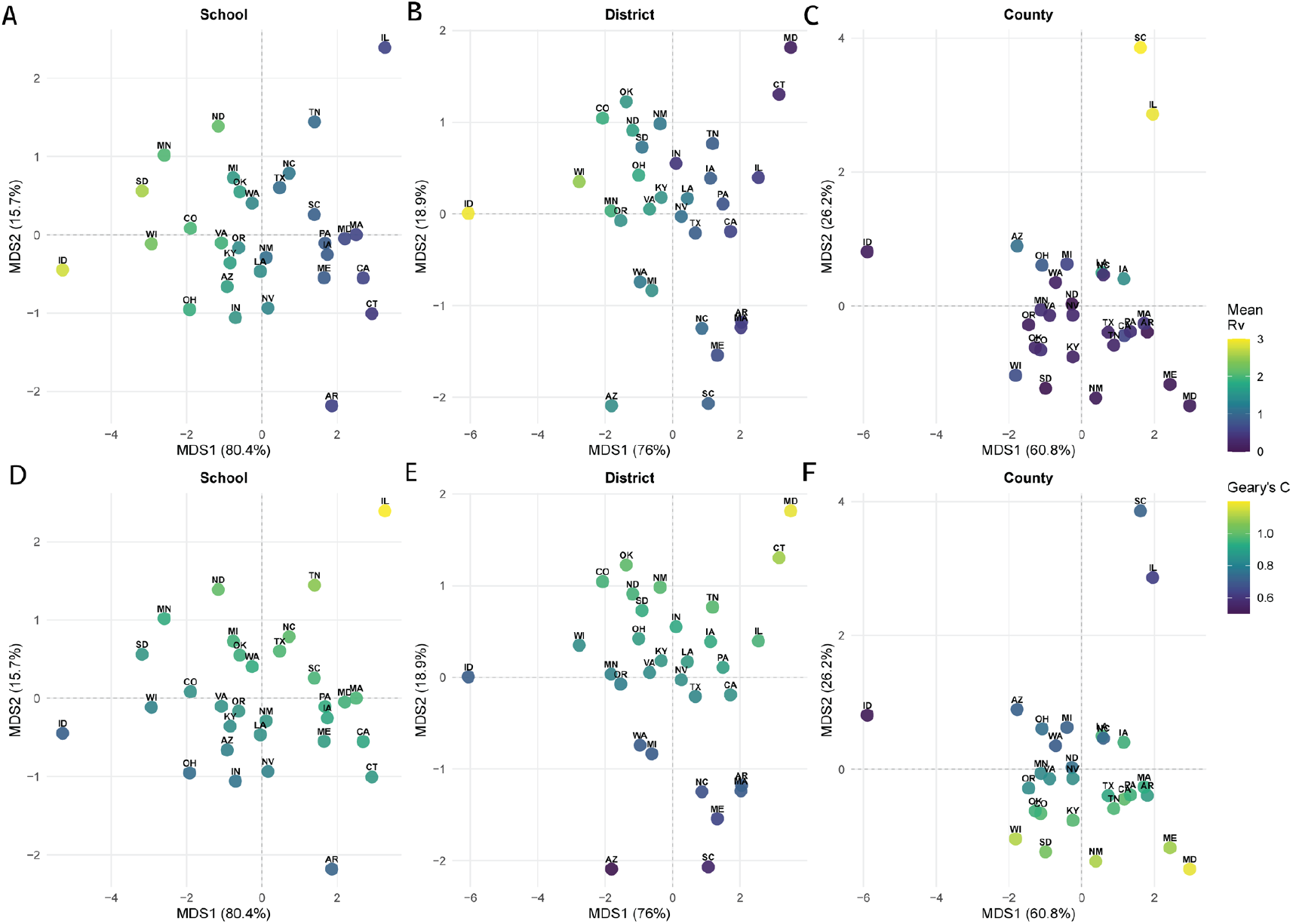
Multi-dimensional scaling of evaluating school-level transmission potential and spatial heterogeneity of susceptibility by state in 2024-2025 school year (or otherwise 2023-2024 school year). 31 states that report MMR vaccination coverage data at all three levels for the 2024-2025 school year (or otherwise 2023-2024 school year) are displayed in the maps. (A-C) dimension1 and 2 colored by mean *R*_*v*_ at the school level, school district level and county levels respectively. (D-F) dimension1 and 2 colored by Geary’s C at the school level, school district level and county level respectively.

The positions of the individual states on the multidimensional scale are loosely constrained by the two-dimensional projection, so two states may be relatively close at the district scale yet much more distant at the county scale. Nonetheless, several robust clusters of states co-occur close together at the school scale and remain grouped as the data are aggregated to district and county levels (Figure 4). States within each cluster share similar coverage levels and heterogeneity, as visible in Figure 1 where they show comparable color intensity and “patchiness”. For example, Iowa, Louisiana and Pennsylvania, cluster closely across all three scales (Figure 4), and display similar depth of color and scattered pockets of lower coverage (Figure 1). Likewise, Kentucky, Minnesota, Oregon, Virginia and Washington cluster more tightly together than Arkansas, Indiana, Illinois and Maine. Understanding the interplay between the level and heterogeneity of coverage is thus a key to interpreting the spatial risk patterns apparent in the maps (Figure 1).

### Reconstructing the accumulated susceptible population

To characterize the age structure and spatiotemporal dynamics of measles susceptibility, we reconstructed the accumulated unvaccinated population. We compiled vaccination records across consecutive kindergarten and 6^th^ or 7^th^ grade cohorts at the county level in six states (Figure 5). Arizona, California, Connecticut, Massachusetts, Minnesota, and Virginia exhibited strikingly divergent susceptibility trajectories (Figure 5), shaped by local policy, demographics, and public health infrastructure.

**Figure 5.**
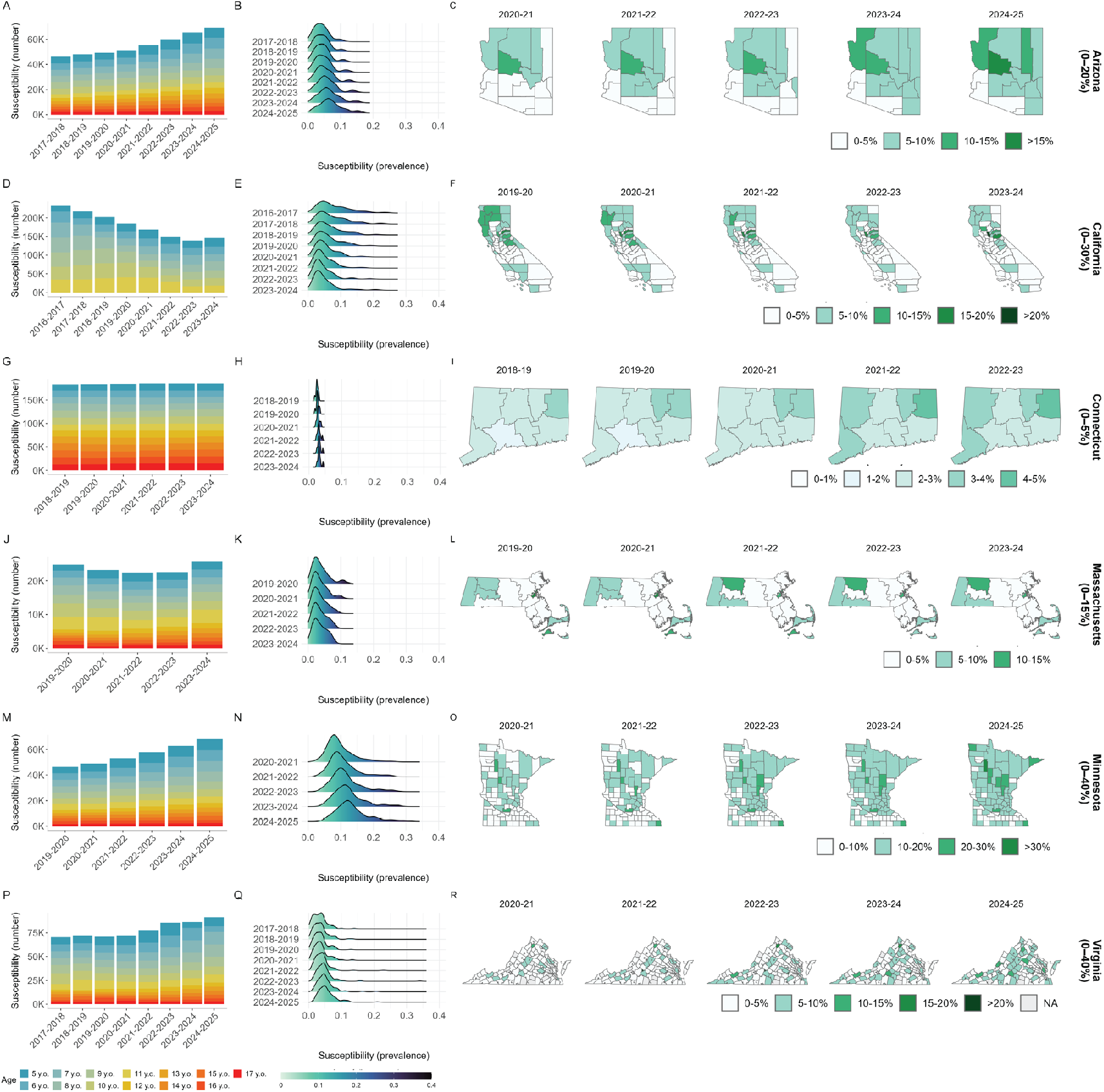
Spatiotemporal dynamics of measles susceptibility reconstructed from time-series MMR vaccination coverage of kindergarten cohorts and 6^th^/7^th^ grade cohorts by county in six states. For Arizona (A–C), California (D–F), Connecticut (G–I), Massachusetts (J–L), Minnesota (M–O) and Virginia (P-Q): (A, D, G, J, M and P) Cumulative number of unvaccinated kindergarteners and 6^th^/7^th^ grades by birth cohort, with colors representing distinct groups currently aged 5–17 years in the 2024-2025 school year (5-12 years in California due to data availability). Each bar represents a school year, and stacked colors indicate the contribution of each birth cohort to the current susceptible population. Ridge plots in (B, H, K, N and Q) show the distribution of prevalence of MMR-susceptible population aged from 5 to 17 years across counties for each available year and from 5 to 12 years in California due to data availability (E). Choropleth maps in (C, F, I, L, O and R) depict county-level susceptibility across selected years, with darker green indicating a higher prevalence of measles susceptibility. Grey areas indicate missing data.

Left-hand panels display the cumulative buildup of unvaccinated individuals aged 5–17 years by state. Arizona displayed intermediate patterns with moderate susceptible accumulation (Figure 5A), ridge plots showing moderate prevalence distributions with substantial inter-annual variability (Figure 5B) and persistent regional clustering (Figure 5C). California displayed a distinct pattern: following implementation of SB277 in 2016 [18], which eliminated non-medical vaccination exemptions, the accumulated susceptible population declined as older under-vaccinated cohorts aged out and were replaced by younger cohorts with higher coverage (Figure 5D). This policy effect is visible in the ridge plots which show a leftward shift, and the geographic mapping, showing the proportion of the population aged 5–17 years who remain unvaccinated in each county (Figure 5E-F). Connecticut maintained low overall susceptible accumulation (Figure 5G), with ridge plots showing stable, low-prevalence distributions (Figure 5H), though some spatial expansion of under-vaccination is visible in recent years (Figure 5I).

Massachusetts demonstrated low, stable susceptible accumulation (Figure 5J), with narrow ridge plot distributions showing minimal inter-annual variation (Figure 5K) and geographic mapping confirming consistently low susceptibility across most counties (Figure 5L). Minnesota exhibited the most concerning pattern, with the highest susceptibility prevalence among all the states examined and substantial accumulation across birth cohorts (Figure 5M). Ridge plots revealed broad, high-prevalence distributions extending well beyond other states, with pronounced rightward temporal shifts indicating worsening trends (Figure 5N). Geographic patterns showed widespread heterogeneity, with hotspots initially concentrated in central Minnesota around 2016– 2017, subsequently spreading throughout the state to create multiple high-susceptibility regions (Figure 5O).

Virginia maintained low susceptible populations with stable (Figure 5P), narrow distributions that kept the majority of counties above the herd immunity threshold; however, in recent years, southwestern counties began accumulating susceptible individuals, as evidenced by an emergent long tail in the ridge plots (Figure 5Q) and substantial spatial spreading of under-vaccination over the past decade (Figure 5R).

These divergent state trajectories: California’s improvement, Minnesota’s deterioration, Massachusetts and Virginia’s emerging vulnerability, and Arizona’s persistent heterogeneity demonstrate that measles susceptibility is neither uniformly increasing nor decreasing nationally but rather follows state-specific dynamics shaped by local policy, demographics, and public health infrastructure.

### Spatial mixing creates transmission risk spillovers

We developed a multiscale framework to estimate county-level effective reproduction numbers that accounts for within-county vaccination heterogeneity and cross-boundary transmission. As shown in Figure 5 and Figure S7, susceptible school-aged children (ages 5–17) are not uniformly distributed but instead cluster spatially within counties. Moreover, school district boundaries frequently do not align with county boundaries (Figure 6A-B), creating opportunities for transmission across administrative units when children from neighboring counties attend the same schools or share community spaces. To translate this fine-scale heterogeneity into county-level outbreak risk, we constructed an age-by-place mixing matrix. This matrix combines an empirical age-specific contact matrix [19] with a spatial connectivity masking matrix derived from school district geography (Figure 6 C-D). The spatial connectivity masking matrix incorporates spillover effects through overlapping catchment areas illustrated as region A1 in Figure 6B, where districts span multiple counties.

**Figure 6.**
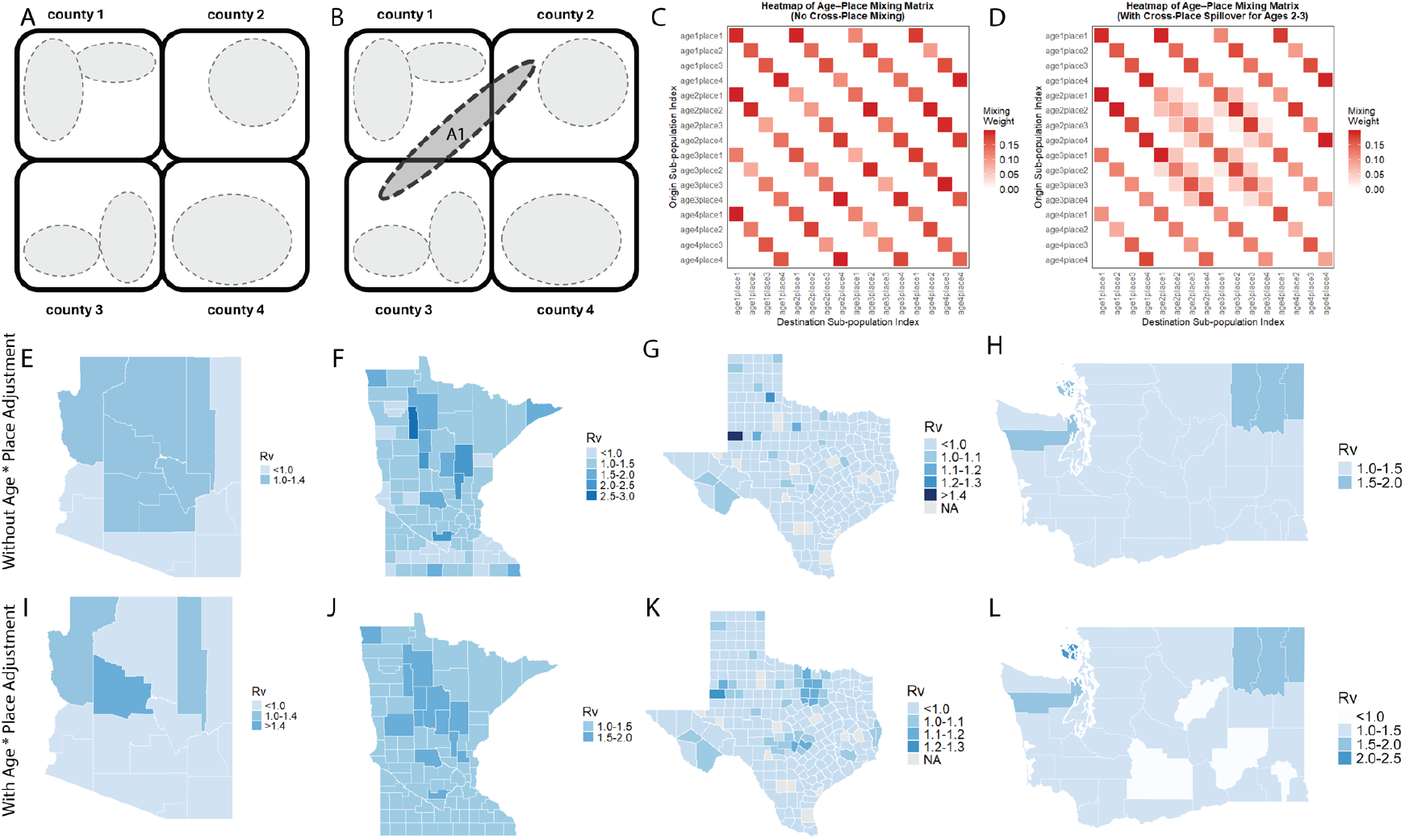
Spatial spillover framework and its impact on county-level measles transmission potential across four US states. (A-B) Conceptual model showing nested county-school district structure: (A) shows school districts (gray ovals) contained within county boundaries with no cross-county mixing, while (B) demonstrates how a multi-county school district (A1, hatched) enables children from counties 1, 2 and 3 to mix across administrative boundaries. (C) and (D) display the corresponding age-place mixing matrices, where (C) shows the block-diagonal structure when mixing occurs only within counties, and (D) shows the emergence of off-diagonal elements when cross-county spillover among school-age children (ages 5-17) is incorporated. (E, I) Arizona, (F, J) Minnesota, (G, K) Texas, and (H, L) Washington *R*_*v*_ estimates comparing homogeneous mixing assumptions (top) versus age-by-place mixing models incorporating cross-boundary transmission (bottom). Blue shading intensity corresponds to *R*_*v*_ magnitude, with darker colors indicating higher transmission potential.

Under the conventional assumption of homogeneous mixing within counties, *R*_*v*_ estimates appear relatively uniform across space (Figure 6E-H). However, when age-place structured mixing is incorporated, *R*_*v*_ estimates reveal substantially greater spatial heterogeneity with localized hotspots exceeding epidemic threshold (Figure 6I-L). These elevated values reflect the combined effect of clustered susceptibility among school-aged children and cross-boundary transmission spillover pathways that link communities across county lines. Together, these findings demonstrate that traditional county-level surveillance metrics may substantially underestimate localized outbreak risk not only by obscuring both the fine-scale spatial structure of immunity gaps and the role of mobility in connecting vulnerable populations, but also by missing how even communities with coverage only modestly below herd immunity thresholds face amplified risk when susceptibles cluster spatially.

## Discussion

The United States crossed the epidemic threshold since 2022: average school-level effective reproduction numbers (*R*_*v*_) exceeded 1.0, creating transmission conditions that support sustained measles outbreaks. This transition, steadily rising since 2013 and doubling post-COVID from ∼5% to 8–10% susceptibility amid worsening spatial clustering, remains invisible to county-level surveillance reporting reassuring averages below threshold throughout. Three mechanisms together explain this aggregation bias. First, enrollment-weighted averaging dilutes small under-vaccinated schools into larger administrative units with higher coverage. Second, distance-driven attenuation inherent to gravity-based mixing kernels reduces *R*_*v*_ estimates at coarser spatial resolutions. Third, aggregation erases the clustering signal detectable through spatial autocorrelation, transforming heterogeneous risk landscapes into falsely uniform surfaces. The magnitude of this bias is substantial: at comparable average susceptibility levels (∼7–8%), school-level *R*_*v*_ values span 0.5–2.5 while county-level values compress below 1.0.

These patterns, long theorized [10] but never empirically validated across nationwide multiscale data, emerge clearly from our assembly of vaccination records spanning 45 states and 50,000+ schools. Progressive disaggregation from county to district to school exposed transmission-permissive clusters invisible at coarser resolutions. Recent county-level analyses [8-9] reveal declining MMR coverage invisible at state scales. Our school- and district-level analysis of the same data uncovers transmission-permissive clusters with *R*_*v*_ >1.0 that county averaging conceals, demonstrating that even improved county resolution remains insufficient for outbreak prediction.

Quantifying the spatial structure of outbreak vulnerability in the absence of circulating pathogen is a fundamental challenge in post-elimination epidemiology. Our findings confirm that pre-elimination and post-elimination settings exhibit mirror-image spatial structure. Classic studies characterized how spatial coupling, hierarchical spread from urban centers and lateral exchange among neighboring towns, shaped fadeout-reintroduction cycles during the transition toward elimination [14-15, 20]. We demonstrate that analogous architecture governs the reverse transition: gravity-based connectivity shapes not only how infection spreads but how susceptibility accumulates. The risk synchrony we observe that clustered vulnerability rising in concert across connected communities is the post-elimination analog to the transmission synchrony documented in endemic metapopulations. This symmetry suggests that theoretical frameworks developed to understand measles persistence can be repurposed to anticipate outbreak vulnerability before cases occur, transforming reactive outbreak response into proactive risk detection.

School districts, not counties, represent the functionally connected populations across which measles can propagate in the United States. Districts frequently span multiple counties while single counties contain portions of many districts. Three spatial phenomena emerged from incorporating this cross-boundary structure into our transmission model. First, connectivity through shared districts increased local effective reproduction numbers across all populations examined, consistent with metapopulation theory predicting elevated persistence under high dispersal. Second, we identified an immunity paradox analogous to source–sink dynamics: well-vaccinated counties functioned as sinks, receiving disproportionate transmission pressure from less-vaccinated source populations. Several such counties crossed the epidemic threshold solely through spillover, despite local immunity levels that would prevent invasion under isolation. Third, central counties exhibited transmission potential exceeding any individual neighboring county, an emergent property of simultaneous connectivity to multiple source populations. This immunity paradox inverts conventional assumptions about local preparedness that a county health department observing reassured local coverage may remain vulnerable to epidemic invasion through pathways its surveillance cannot see. The spatial grain of management interventions must match the scale of transmission to effectively prevent outbreaks.

State trajectories expose intervention windows. California’s SB277 bill in 2016 produced measurable improvement as under-vaccinated cohorts aged out, demonstrating that policy intervention can reverse susceptible accumulation. Minnesota’s continued deterioration and Virginia’s emerging vulnerability illustrate the alternative trajectory when intervention is absent. These state-specific dynamics are shaped by exemption policies, demographic structure, and public health infrastructure demand surveillance systems capable of detecting local trends rather than relying on national averages [21-22]. More broadly, the post-COVID-19 doubling of susceptibility from approximately 5% to 10% reflects a global phenomenon: Europe reported its highest measles incidence in 25 years, Canada lost elimination status, and the United Kingdom recorded its first child death in a decade. The return of measles in high-income countries stems not from vaccine failure but surveillance granularity mismatched to transmission scales.

Several limitations warrant consideration. Our analysis relies on kindergarten vaccination records as a proxy for cumulative susceptibility; delayed vaccination or interstate migration may introduce misclassification. The gravity-based *R*_*v*_ formulation assumes mixing proportional to distance and enrollment but does not incorporate detailed mobility data, which remain sparse for school-aged populations. State-level data heterogeneity, varying completeness, exemption reporting, and temporal coverage, introduces uncertainty in cross-state comparisons. Finally, our framework quantifies transmission potential rather than realized outbreak risk, which depends additionally on importation frequency, and stochastic dynamics.

Measles elimination across the Americas required decades of sustained vaccination effort. Our findings reveal that this achievement is now structurally vulnerable: transmission potential has crossed the epidemic threshold at the scale where outbreaks originate, susceptible populations continue growing in spatially clustered pockets connected through shared school infrastructure, and surveillance systems designed for aggregated monitoring cannot see the danger of building. The spatial grain of management interventions must match the scale of transmission to effectively prevent outbreaks. These findings come at a critical moment: with 2025 US cases exceeding 2,000 (highest in 30 years). District-county misalignment creates invisible spillover pathways. Averting endemic re-establishment demands real-time school and district dashboards, cross-jurisdictional coordination, and targeted campaigns in vulnerability clusters. Reversing courses are possible but requires surveillance that sees risk where it exists: at the school and district level, before imported cases ignite community chains.

## Material and Methods

### Data Sources and Acquisition

#### Vaccination data

Immunization data were obtained from state health departments for kindergarten and, where available, 6^th^ and 7^th^ grade entrance surveys and Washington Post publicly available datasets [23]. Data included Measles-Mumps-Rubella (MMR) vaccination coverage and school enrollment at the school, school district, and county levels when reported. Data availability varied by state and year; a comprehensive summary of data sources, years of coverage, and reporting formats is provided in Table S1.

#### Geocoding

Individual school geographic coordinates (latitude and longitude) were obtained using the Google Cloud Geocoding API based on school names and addresses.

#### Administrative boundaries

School district boundaries were obtained from the National Center for Education Statistics (NCES) Education Demographic and Geographic Estimates (EDGE) program [24]. County boundaries were obtained from the U.S. Census Bureau’s TIGER/Line Shapefiles [25].

#### Data Processing and Aggregation

For states where school district- and county-level MMR coverage estimates were not directly reported, we aggregated school-level data using the following approach: (1) geocoded all schools to obtain point locations; (2) performed spatial joins to assign each school to its corresponding district and county based on administrative boundary shapefiles; and (3) calculated district- and county-level coverage as the enrollment-weighted mean of school-level coverage:

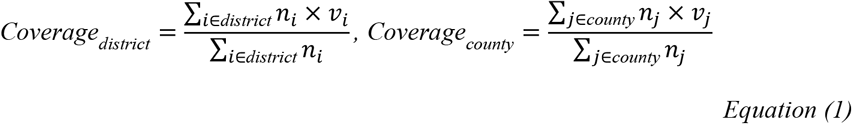

where *n*_*i*_ and *n*_*j*_ is the enrollment and *v*_*i*_ and *v*_*j*_ is the vaccination coverage at school *i* and *j*. Note that private schools were included in school district- and county-level aggregations, although private schools do not belong to public school districts in administrative terms. This approach provides a more complete estimate of population-level immunity and susceptibility within geographic areas.

## Metrics and Indices

### Spatial autocorrelation (Geary’s C)

To quantify spatial clustering of susceptibility within states, we calculated Geary’s C statistic for each state-year-scale combination:

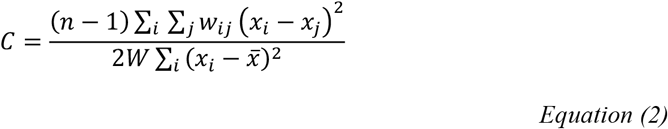

where *x*_*i*_ is the susceptibility (100% − MMR coverage) at unit *i, w*_*ij*_ is the spatial weight between units *i* and *j* (defined as 1 for adjacent units, 0 otherwise), *W* = ∑_*i*_ ∑_*j*_ *w*_*ij*_ is the sum of all weights, and *n* is the number of spatial units. Values of *C* < 1 indicate positive spatial autocorrelation (clustering of similar values), *C* = 1 indicates spatial randomness, and *C* > 1 indicates spatial dispersion. Geary’s C values are not directly comparable across spatial scales due to differences in the number of units, spatial neighbor structures, and variance compression through aggregation.

The definition of spatial neighbors differed by spatial scale. For polygon-based units (school districts and counties), we used queen contiguity, where *w*_*ij*_ = 1 if units *i* and *j* share a border or corner, and *w*_*ij*_ = 0 otherwise. For point-based units (schools), which lack natural boundaries, we defined neighbors using *k-*nearest neighbors with *k* = 5, where *w*_*ij*_ = 1 if school *j* is among the five geographically closest schools to school *i* based on Euclidean distance, and *w*_*ij*_ = 0 otherwise. This approach ensures that each school has a consistent number of neighbors for spatial autocorrelation calculation, regardless of the density of schools in different regions of the state.

### Coverage inequality (Gini coefficient)

To quantify inequality in vaccination coverage within states, we calculated the Gini coefficient for each state-year-scale combination:

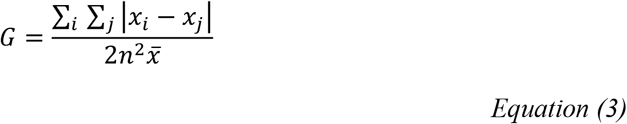

where *x*_*i*_ is the coverage at unit *i, n* is the number of units, and 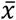 is the mean coverage. Values range from 0 (perfect equality; all units have identical coverage) to 1 (perfect inequality). Gini coefficients are not directly comparable across spatial scales because aggregation compresses the coverage distribution, systematically reducing observable inequality at coarser resolutions.

### Vaccination-adjusted effective reproduction number, *R*_*v*_using a gravity-based mixing kernel

We estimated vaccination-adjusted effective reproduction numbers, *R*_*v*_ at the individual school level, school district level, and county level using a gravity-based mixing kernel adapted from metapopulation transmission theory. Following established frameworks for modeling spatially heterogeneous vaccination coverage and non-random mixing patterns [13], we divided the population into *n* spatial subunits (schools, districts, or counties), each characterized by its own vaccination coverage and spatial connectivity.

#### Next-generation matrix

The effective reproduction number in a spatially heterogeneous population was calculated using the next-generation matrix (NGM) approach. Following standard metapopulation frameworks, we defined the transmission kernel between locations *i* and *j* as:

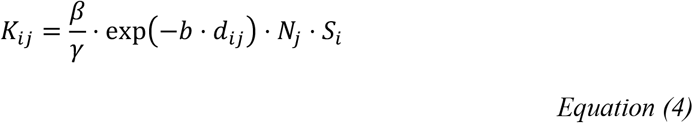

where *d*_*ij*_ is the Euclidean distance (km) between unit centroids calculated using Haversine formula, b = 0.001 km^−1^ is the distance decay parameter [13], *N*_*j*_ is the population (enrollment) at location *j*, and *S*_*j*_ = 1 − *p*_*j*_ is the susceptible fraction at location *j* (with *p*_*j*_ being vaccination coverage). The exponential distance decay function exp−*b* ⋅ *d*_*ij*_ represents the probability of contact between individuals at distance *d*_*ij*_, reflecting empirical patterns of human mobility where contact rates decline with increasing distance.

#### Calibration

The transmission coefficient *β*/*γ* was calibrated such that the spectral radius of the contact matrix equals the basic reproduction number for measles (*R*_0_ = 15) in a fully susceptible population:

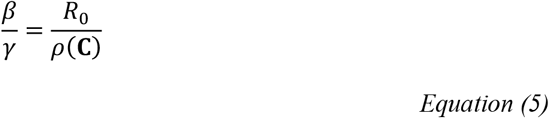

where *ρ*(***C***) is the spectral radius of the contact matrix ***C*** with elements *C*_*ij*_ = exp−*b* ⋅ *d*_*ij*_ ⋅ *N*_*j*_ ⋅ *S*_*i*_. where ***C*** is the contact matrix with elements *C*_*ij*_ = exp(−*b* ⋅ *d*_*ij*_) ⋅ *N*_*j*_, and *ρ*(⋅) denotes the spectral radius (dominant eigenvalue). This calibration ensures that *R*_*v*_ reflects reductions in transmission potential due to vaccination while maintaining consistency with measles epidemiology.

The location-specific effective reproduction number was calculated as the column sum of the NGM:

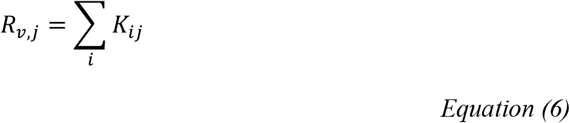

representing the expected number of secondary infections generated by an infected individual at location *j* across all other locations, weighted by distance-based contact probability and local susceptibility. The global Rv for the entire metapopulation was computed as the spectral radius of ***K***: *R*_*v global*_ = *ρ*(***K***).

#### Scale-specific computation

Computational details. Distance matrices were computed using Haversine distances between unit centroids. For school and district levels, the NGM was computed within each state, yielding state-specific *R*_*v*_ estimates. For county level, a single national NGM was constructed with dimension equal to the total number of counties across all states with available data. The spectral radius was computed using standard eigenvalue decomposition for matrices with fewer than 500 rows, and iterative methods (Arnoldi algorithm via RSpectra) for larger matrices.

At the school level, *R*_*v*_ was computed separately within each state to reflect state-level policy environments and maintain computational tractability. At the district level, *R*_*v*_ was similarly computed within states. At the county level, *R*_*v*_ was computed nationally to capture cross-border transmission dynamics, as county-level transmission is more likely to span state boundaries.

### Validation: 2024–2025 Texas Outbreak

To validate our *R*_*v*_ estimates, we compared model predictions with observed transmission dynamics from the 2024–2025 measles outbreak in Gaines County, Texas. From the exponential growth phase of the outbreak (days 5–9), we estimated the exponential growth rate *r* by fitting a linear regression to the natural log of cumulative incidence, yielding *r* = 0.288 per day. The effective reproduction number was then estimated as:

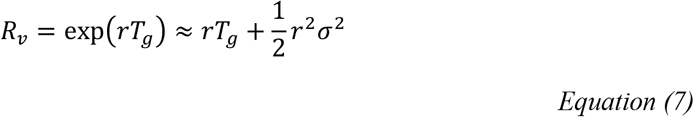

where *T*_*g*_ = 11.1 days is the mean generation time for measles and *σ* = 1.79 days is the standard deviation. This yielded *R*_*v*_ estimates between 3.1 and 3.2. Given the reported vaccination coverage of 70–80% in the affected community, the implied *R*_0_ is approximately 14–16, consistent with our assumed *R*_0_ = 15 for model calibration.

### Estimates for susceptible population aged 5 to 17 years by county

To estimate the prevalence of MMR-susceptible individuals aged 5–17 years at the county level, we combined directly observed vaccination coverage from school entry surveys with cohort-based projections for intervening ages. Kindergarten (age 5–6) and 6th or 7th grade (age 11–13) vaccination coverage was obtained directly from annual state immunization surveys (Table S1). For ages not captured by these surveys, we projected susceptibility by tracking kindergarten and 6^th^ or 7^th^ grade cohorts forward in time, assuming vaccination status at kindergarten entry remained constant until the next survey point at 6th or 7th grade.

For each county *c* and year *t*, the age-specific susceptible population was calculated as:

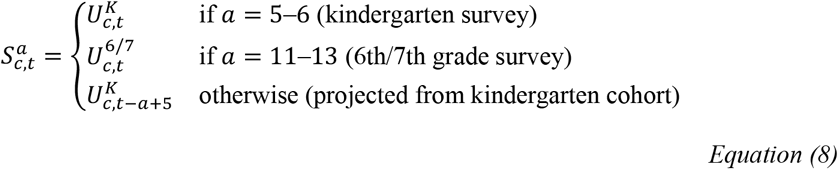

where 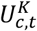 is the number of unvaccinated kindergarteners directly observed in county *c* at year *t*, 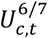 is the number of unvaccinated 6th or 7th graders directly observed, and 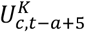 is the projected number of unvaccinated individuals aged *a* based on the kindergarten cohort from year *t* − *a* + 5.

The total susceptible prevalence for ages 5–17 was then calculated as:

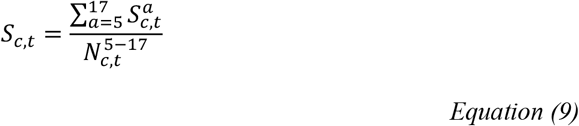

where 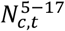 is the total population aged 5–17 years in county *c* at year *t*.

This approach leverages two direct observation points (kindergarten and middle school) while filling gaps through cohort aging (Figure S7). The 6th/7th grade survey provides a validation checkpoint for cohort projections and captures any catch-up vaccination that occurred between kindergarten and middle school entry. However, the projections for non-survey ages do not account for migration or vaccination changes between survey points, although annual intercounty migration among school-age children in the United States is relatively low, typically affecting fewer than 4% of the population (U.S. Census Bureau, 2023) [26].

### Age-by-place multiscale modelling approach

To account for both age-structured contact patterns and spatial heterogeneity in mixing patterns, we developed an age-by-place mixing contact matrix. The model represents the population as a graph where each node corresponds to an (age *a*, county *c*) stratum. Edges encode mixing patterns across ages (based on empirical contact matrices) and between places (based on a gravity model of spatial connectivity). County-level effective reproduction numbers are derived from the spectral properties of the resulting next-generation matrix.

The age-specific contact matrix *C*_*age*_ is a 105-by-105 matrix obtained from [ref], where elements *C*_*age*_[*i, j*] represents daily contact between individuals of age *i* and age *j*. We define the county adjacency matrix *A*_*county*_ as an *n*_*counties*_ × *n*_*counties*_ binary matrix:

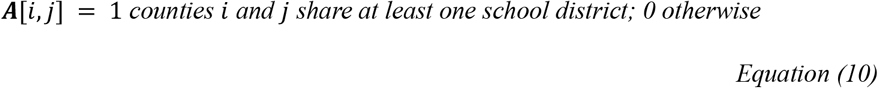

with diagonal elements set to zero to exclude within-county transmission from the cross-county component.

The total mixing matrix ***M***_*total*_ has dimension (*n*_*ages*_ × *n*_*counties*_) × (*n*_*ages*_ × *n*_*counties*_) and comprises two components. The within-county component allows all age groups to mix according to the full contact matrix:

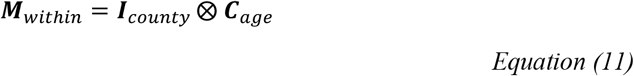

In the cross-county mixing component, only children (ages 5–17) mix across counties via shared school districts:

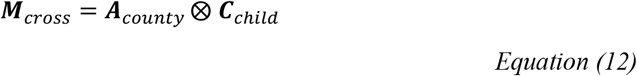

Where ***C***_*child*_ retains only the 5-17 age block of ***C***_*age*_ and sets all other elements to zero. The total mixing matrix is then:

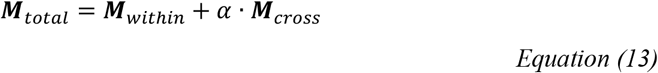

where *α* is a spillover strength parameter calibrated separately for each state (fixed at 0.1 for all states examined, including Texas, Minnesota, Arizona and Washington).

To incorporate vaccination coverage, we define a susceptibility vector ***s*** with elements,

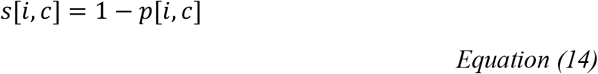

where, *p*[*i, c*] is the vaccination coverage for age *i* in county *c*. School-age coverage (ages 5-17) was obtained from state immunization records; coverage for other age groups was imputed based on CDC estimates (90% ages 1-4; 95% for ages 18+) [ref]. The diagonal susceptibility matrix is

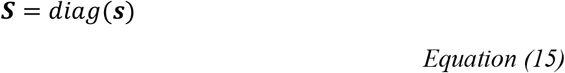

The next generation matrix is

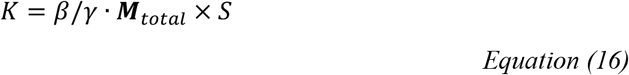

where *β*/*γ* is scales such that the dominant eigenvalue of ***K*** equals the basic reproduction numbers *R*_0_ = 15 for measles in a fully susceptible population. Element ***K***[*i, j*] represents the expected number of secondary infections in group *i* generated by a single infected individual in group *j*.

To obtain county-level reproduction numbers, we aggregate across age strata using the mapping matrix ***G*** of dimension (*n*_*ages*_ × *n*_*counties*_) × *n*_*counties*_, where *G*[*i, c*] = 1 if index *I* belongs to county *c* and 0 otherwise. The county-level NGM is

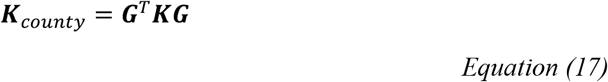

The effective reproduction number of county *c* is the corresponding row sum:

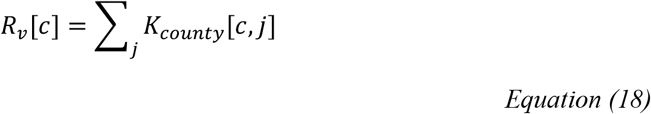

This quantity can be decomposed into within-county and spillover components by applying the same aggregation separately to

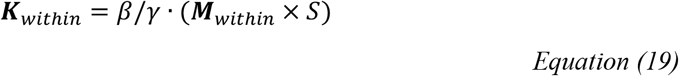

and

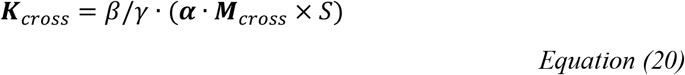

yielding 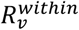 and 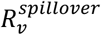 such that:

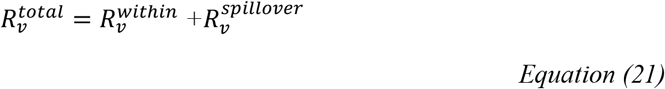

## Data and Code Availability

All code and data will be available upon submission to a peer reviewed journal

## Acknowledgements

We thank the Bento lab for comments on this work.

## Funding

Cornell Atkinson Center seed funds.

## Author contributions

Conceptualization: S.C., A.B. Methodology: S.C., A.B. Investigation: S.C., Visualization: S.C., Funding acquisition: A.B.. Project administration: A.B. Supervision: A.B. Writing – review & editing: S.C., A.B.

## Competing interests

The authors declare that they have no competing interests.

## Supplementary Information

**Figure S1.**
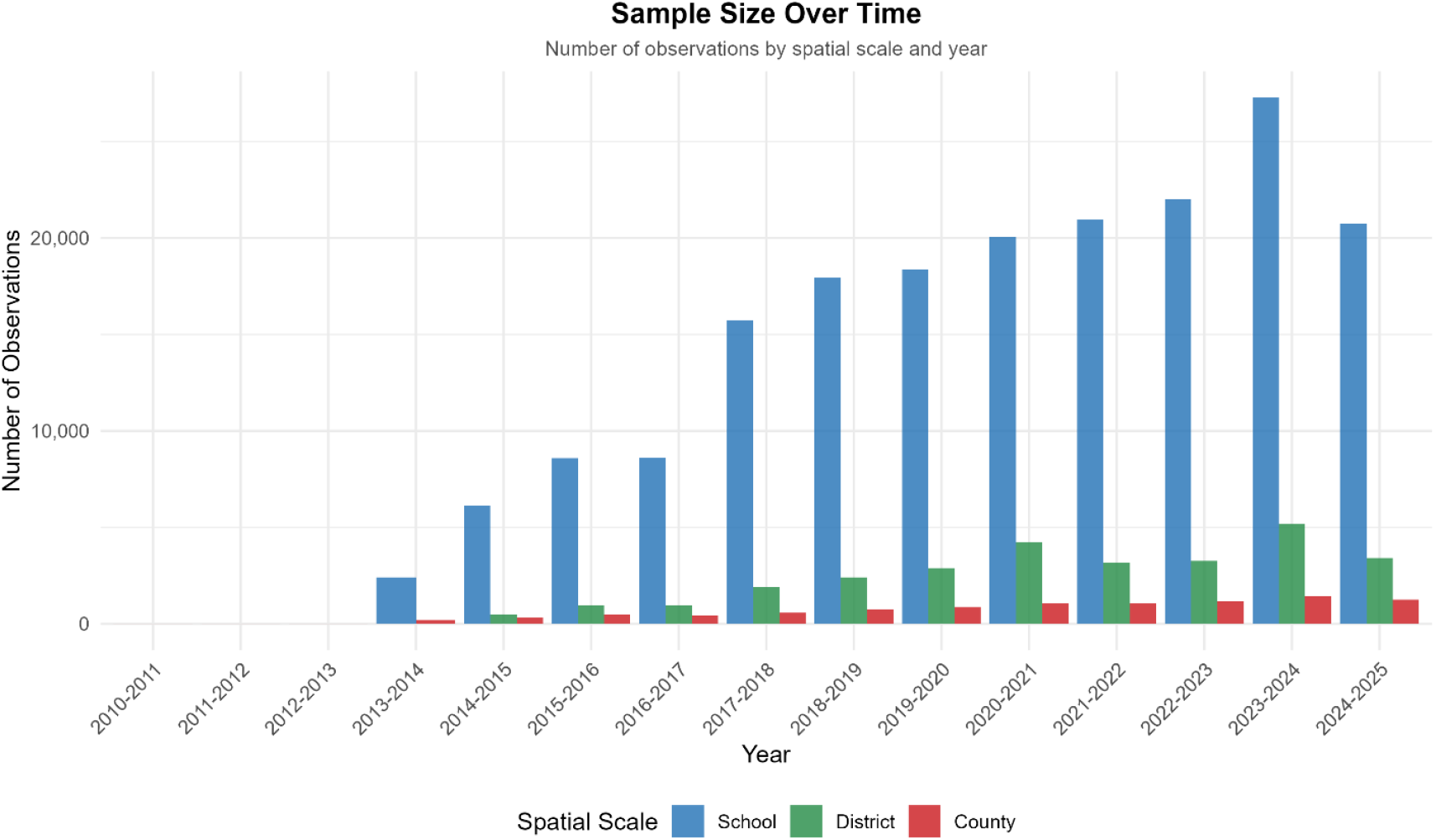
Sample size by spatial scale and school year. Number of observations included in the analysis at individual school (blue), school district (green), and county (red) levels from 2013–2025.

**Figure S2.**
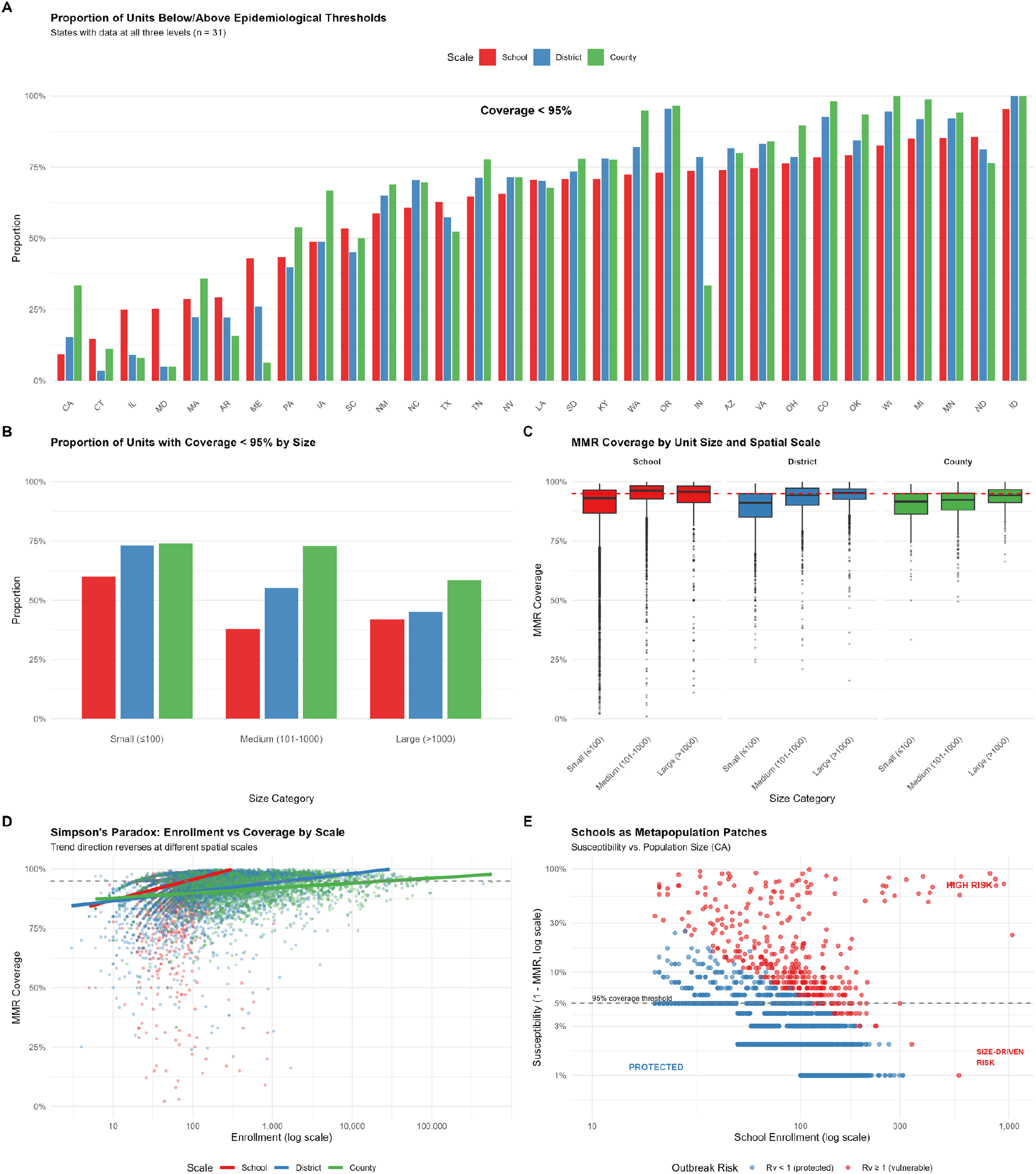
Aggregation bias and Simpson’s paradox in vaccination coverage across spatial scales. (**A**) Proportion of units with MMR coverage below the 95% herd immunity threshold at school (red), district (blue), and county (green) levels across 31 US states. States are ordered by school-level proportion below threshold. Paradoxically, in many states (particularly those with higher overall coverage on the right), a higher proportion of counties fall below the 95% threshold than individual schools. This counterintuitive pattern arises because large schools have higher coverage than small schools, and enrollment-weighted aggregation dilutes the contribution of small, under-vaccinated schools. The result is that county-level averages can fall below threshold even when the majority of individual schools exceed it—demonstrating how aggregation can either mask or amplify apparent vulnerability depending on the size-coverage relationship. (B) Proportion of units below 95% coverage stratified by enrollment size category. At the school level (red), small units (≤100 enrollment) show the highest proportion below threshold, while large schools (>1000) show the lowest. However, at district (blue) and county (green) levels, this gradient flattens or reverses, reflecting how the amalgamation of heterogeneous schools’ obscures size-dependent coverage patterns. (**C**) MMR coverage distributions by unit size and spatial scale. Within each scale, larger units consistently show higher median coverage and narrower distributions than smaller units. This size-coverage relationship is the mechanistic driver of the aggregation paradox: because large schools vaccinate more effectively, their disproportionate weight in enrollment-based aggregation elevates district and county averages while masking vulnerability concentrated in small schools. (**D**) Simpson’s paradox in the enrollment-coverage relationship. At the school level (red), coverage increases strongly with enrollment— larger schools vaccinate more effectively. This positive relationship attenuates at the district level (blue) and weakens further at the county level (green). The divergent slopes demonstrate how ecological inference from aggregated data can misrepresent unit-level relationships, with implications for targeting interventions based on administrative-level coverage estimates. (**E**) Schools as metapopulation patches: susceptibility versus enrollment for California schools on log-log axes. Points are colored by outbreak risk classification (*R*_*v*_ ≥ 1, vulnerable; *R*_*v*_ < 1, protected). The horizontal dashed line indicates the 5% susceptibility threshold corresponding to 95% coverage. Three distinct zones emerge: (1) protected schools below the threshold with low susceptibility regardless of size, (2) high-risk schools with elevated susceptibility, predominantly small schools with variable coverage, and (3) size-driven risk where large schools with moderate susceptibility cross the epidemic threshold due to enrollment-dependent transmission dynamics in the gravity-based Rv formulation. This metapopulation structure underlies the spatial heterogeneity that aggregated surveillance fails to capture.

Many states show lowest coverage at the county level, in terms of less than 95 % vaccinated, and highest at the school level despite that these are all compiled from the same basic school data, number unvaccinated in the 23-24 exercise versus the number enrolled eligible for vaccination (Figure S2(A)). The difference arises through the pooling of data from all the schools in the district or all the schools in a county. This is an example of Simpson’s paradox.

The Simpson paradox may be understood through the introduction of confounding variables, Figure S2(B) compares the patterns of coverage when the analysis is done separately on small, medium and large schools. This shows that the discrepancy among the school, district and county levels is largely accounted for by school size. Large schools show much higher coverage than small schools, and this is also reflected in the district-level figures for districts containing large schools. Another view of this pattern can be seen in Figure S2(C) where the county level coverage is little affected by school size, compared to the less balanced patterns at the school and district levels. Small schools evidence less coverage especially at the level of district and school. Figure S2(D) shows a regression-based analysis of coverage versus school enrollment, where at each scale larger schools have better coverage, an effect which is more marked at the school level than at the district level, and even more so at the county level. The latter analysis is based on raw school coverage values rather than proportions of schools that fall on each side of a cut-off level of 95%. Nevertheless, the two approaches are confirmatory.

The greater dispersion of coverage values at the school level suggests a closer look at the distribution of the data as in Figure S2(E). We see that as expected, susceptibility is greater at the school level than the county level. For all three levels, school, district and county, susceptibility decreases as enrollment increases, but at the school level, this trend is disrupted by an appreciable number of larger schools which resist the tendency to have less susceptibility.

**Figure S3.**
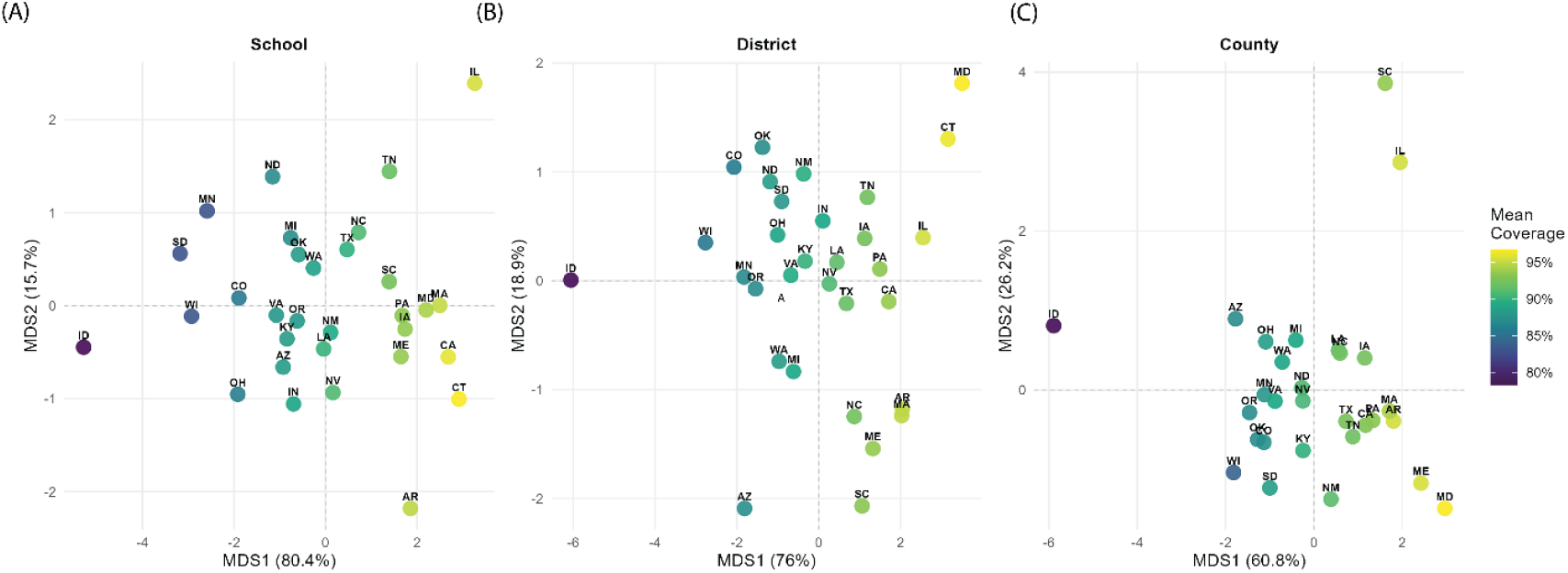
Multi-dimensional scaling of evaluating school, school district and county level transmission potential and spatial heterogeneity of susceptibility by state in 2024-2025 school year (or otherwise 2023-2024 school year). 31 states that report MMR vaccination coverage data at all three levels for the 2024-2025 school year (or otherwise 2023-2024 school year) are displayed in the maps. (A-C) dimension1 and 2 colored by mean MMR vaccination coverage at the school level, school district level and county levels respectively.

**Figure S4.**
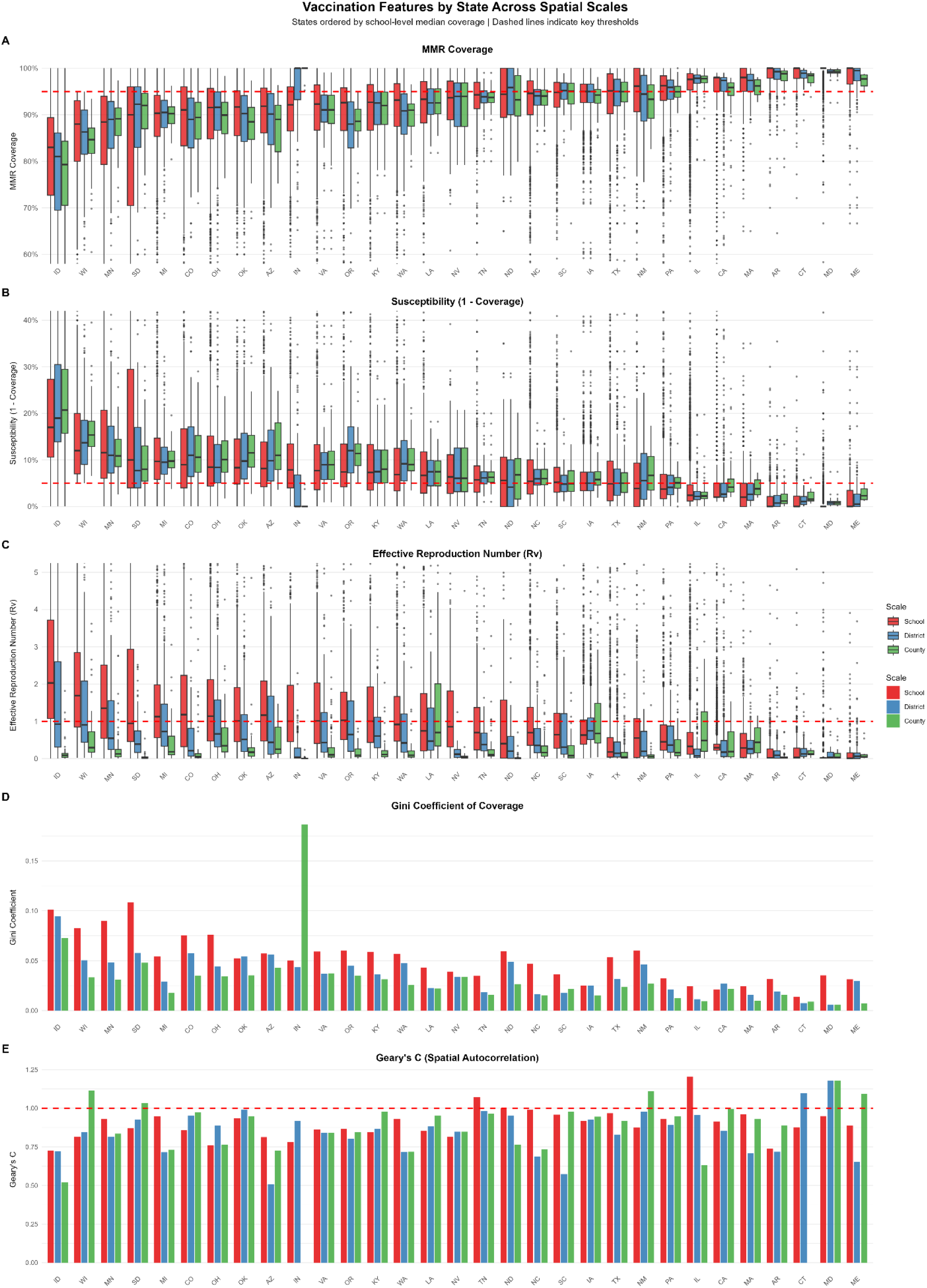
Distribution of vaccination features by state across spatial scales. **(A)** MMR vaccination coverage distributions at school (red), district (blue), and county (green) levels for 31 US states. States are ordered by school-level median coverage from lowest (Idaho) to highest (California). The dashed red line indicates the 95% herd immunity threshold. Coverage distributions are widest at the school level, reflecting substantial within-state heterogeneity that is progressively obscured by aggregation to district and county scales. **(B)** Susceptibility (1 − coverage) distributions across scales. The dashed red line marks the 5% susceptibility threshold corresponding to herd immunity. States with low median coverage (left) show substantial proportions of schools exceeding 20–30% susceptibility, while even high-coverage states contain individual schools with elevated susceptibility. **(C)** Effective reproduction number (Rv) distributions derived from gravity-based transmission models. The dashed red line indicates the epidemic threshold (Rv = 1). School-level Rv distributions consistently exceed district and county levels due to enrollment weighting and distance attenuation effects. Several states show median school-level Rv above 1, indicating transmission-permissive conditions at fine spatial scales despite lower aggregate estimates. **(D)** Gini coefficient of coverage inequality within each state. Higher values indicate greater heterogeneity in vaccination coverage. School-level Gini coefficients (red) consistently exceed district (blue) and county (green) levels, demonstrating that coverage inequality is most pronounced at fine spatial scales. Idaho and Minnesota exhibit the highest school-level inequality. **(E)** Geary’s C statistics measure spatial autocorrelation of susceptibility. Values below 1.0 (dashed line) indicate positive spatial autocorrelation (clustering of similar values); values above 1.0 indicate spatial dispersion. Most states show Geary’s C < 1 at the school level, indicating that under-vaccinated schools tend to cluster spatially rather than distribute randomly—a pattern that amplifies local outbreak risk beyond what random mixing models would predict.

**Figure S5.**
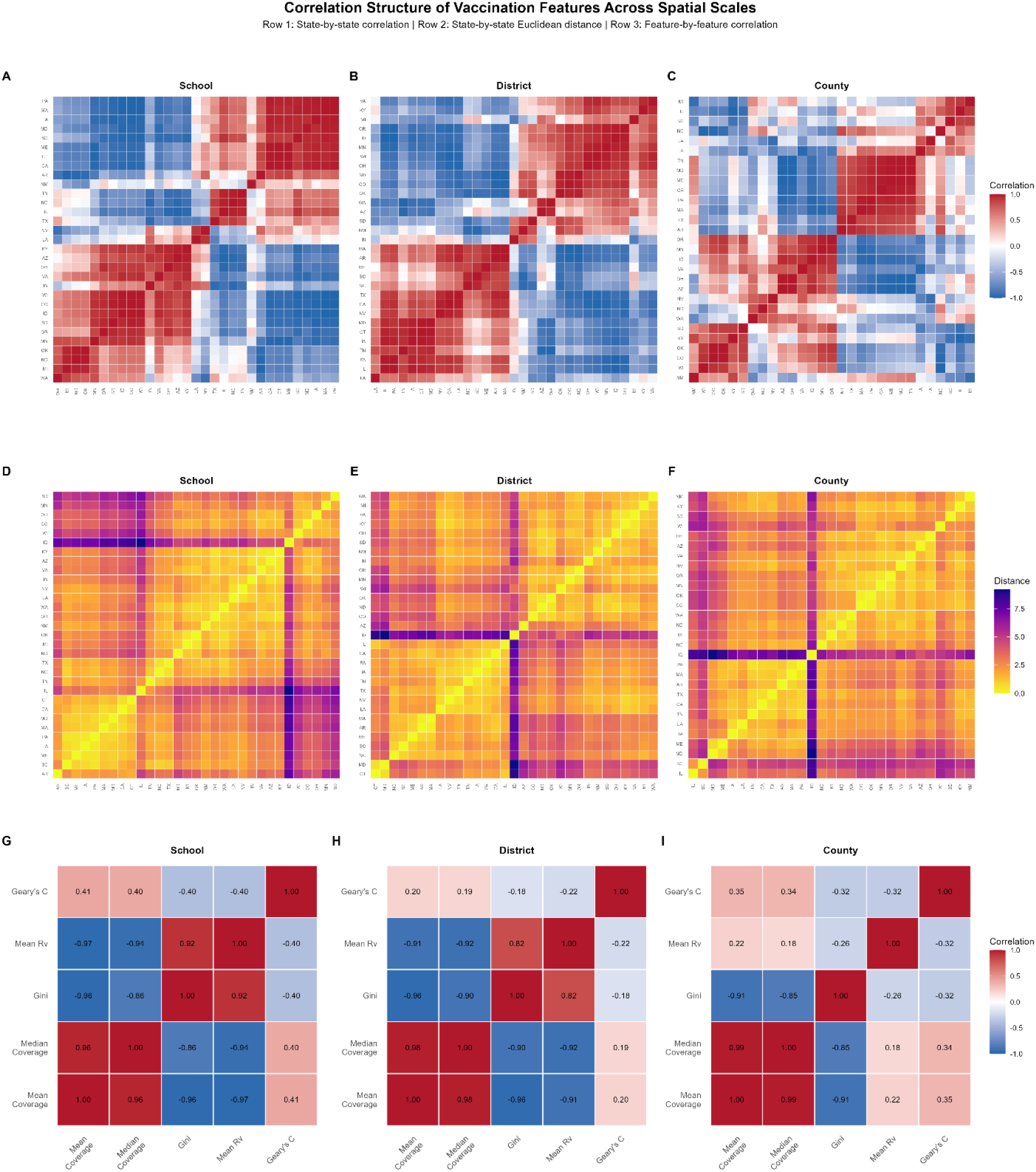
Correlation structure of vaccination features across spatial scales. (A–C) State-by-state Pearson correlation matrices based on standardized vaccination feature profiles (mean coverage, median coverage, Gini coefficient, mean *R*_*v*_, and Geary’s C) at the school (A), district (B), and county (C) levels. States are ordered by hierarchical clustering (Ward’s method), revealing groups with similar vaccination characteristics. Red indicates positive correlation (similar profiles); blue indicates negative correlation (dissimilar profiles). **(D–F)** State-by-state Euclidean distance matrices at school (D), district (E), and county (F) levels. Yellow indicates small distances (similar states); purple indicates large distances (dissimilar states). Clustering structure is largely preserved across scales, though some state groupings shift with aggregation. **(G–I)** Feature-by-feature correlation matrices showing relationships among the five metrics at school (G), district (H), and county (I) levels. Mean and median coverage are strongly positively correlated (r > 0.96) and strongly negatively correlated with Gini coefficient and mean *R*_*v*_ across all scales. Geary’s C shows weaker associations with other features, particularly at the county level, indicating that spatial autocorrelation captures independent information about vaccination landscape structure. The attenuation of correlations from school to county level reflects how aggregation obscures the relationships between coverage heterogeneity and transmission potential.

**Figure S6.**
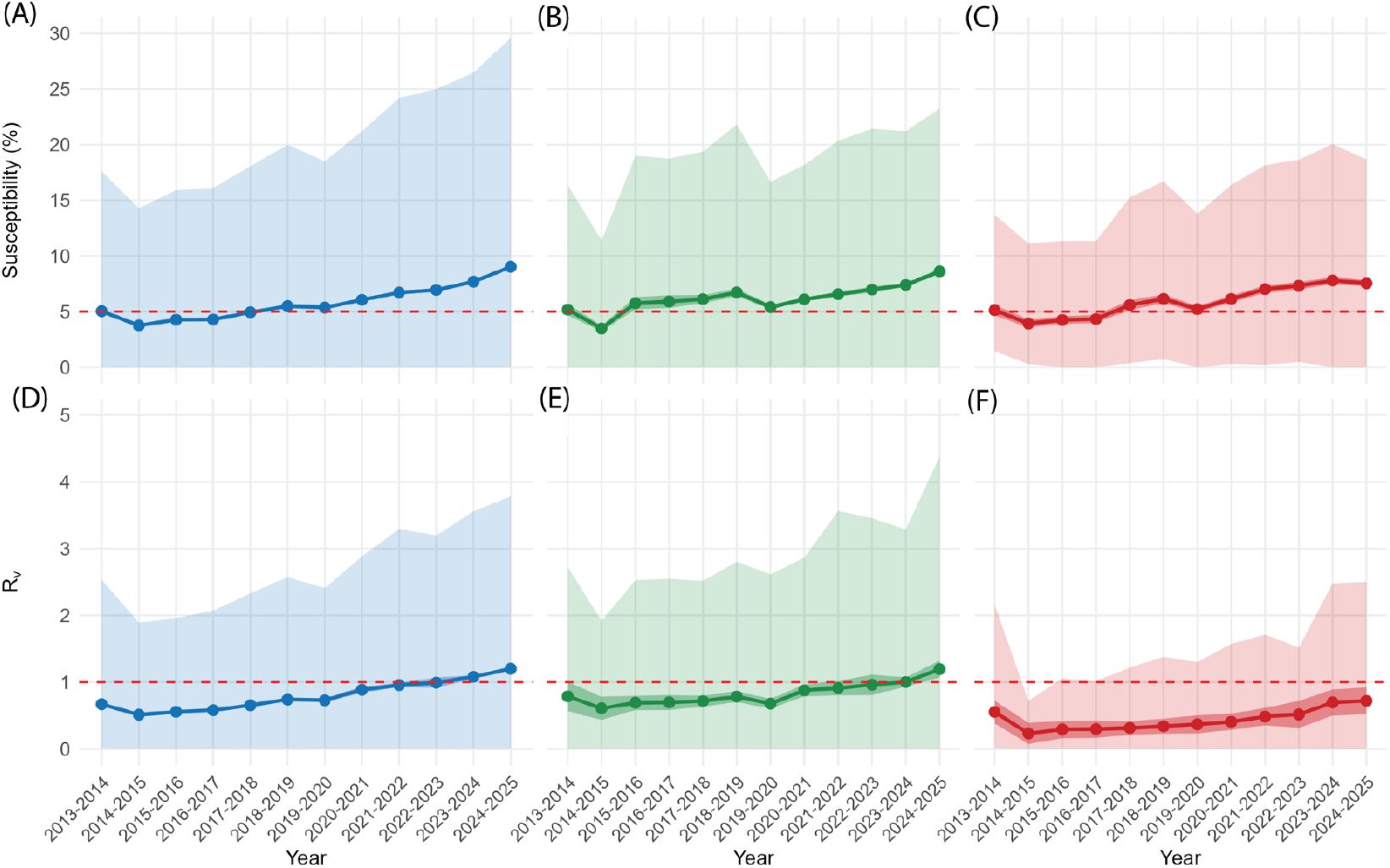
(A–C) Time course of average susceptibility (points and solid lines) with 95% confidence band (central darker shaded ribbons, calibrated with mean ±1.96×standard deviation) and 90% inter-quantile range (IQR) at the individual school, school district, and county level from 2013-2014 to 2024-2025 school year among states. The red horizontal dashed line denotes the susceptible pool threshold, susceptibility = 5%. (D-F) Time course of average *R*_*v*_ (points and solid lines) with 95% confidence band (central darker shaded ribbons, calibrated with mean ±1.96×standard deviation) and 90% inter-quantile range (IQR) at the individual school, school district, and county level from 2013-2014 to 2024-2025 school year among states. The red horizontal dashed line denotes the epidemic threshold, *Rv* = 1.0.

**Figure S7.**
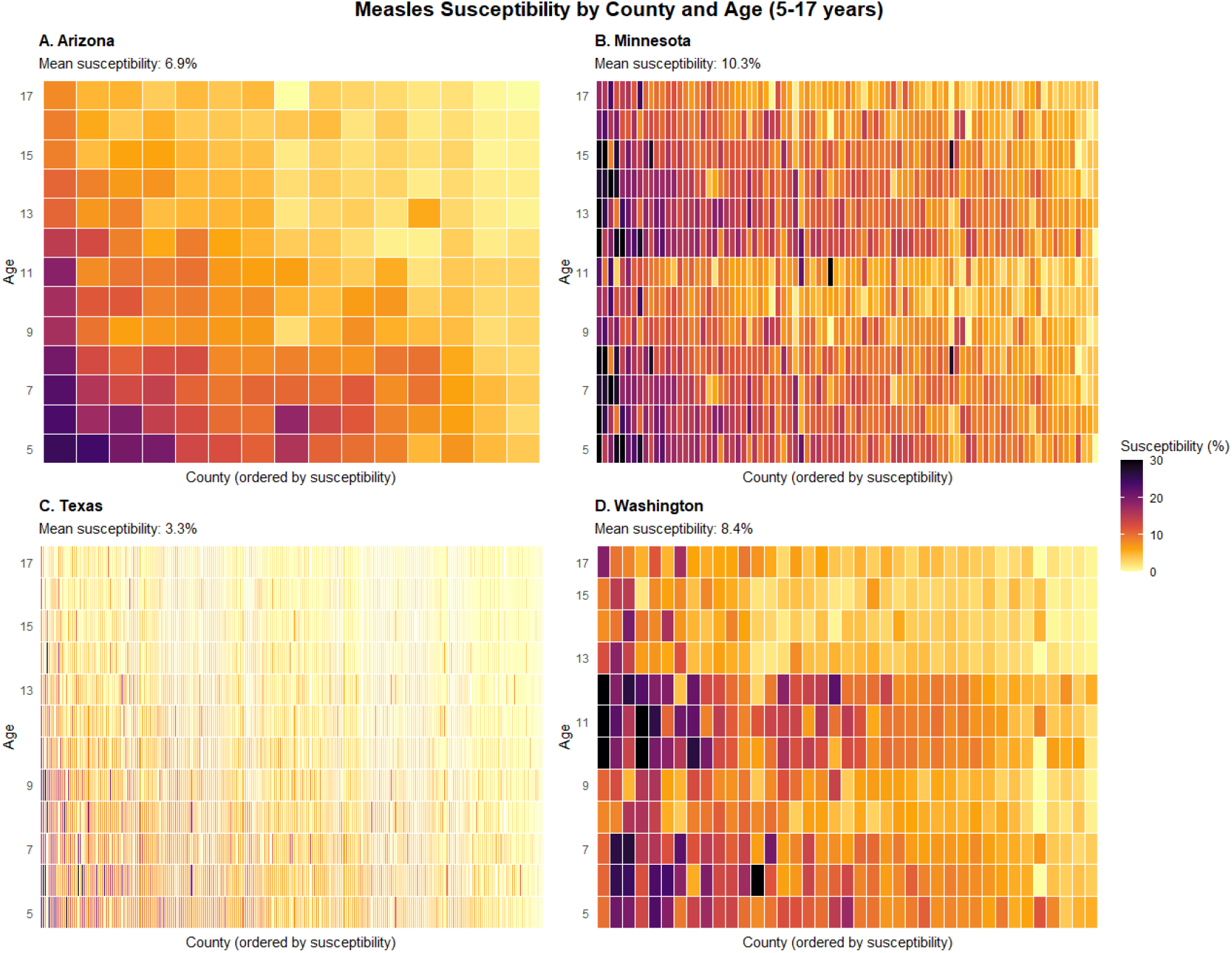
MMR susceptibility (100%-MMR coverage) by county and by age in Arizona (A), Minnesota (B), Texas (C) and Washington (D) in 2024-2025 school year. The MMR coverages by age and by county is estimated through yearly kindergarten entrance survey by county and 6^th^ or 7^th^ grade entrance immunization survey by county from 2013-2024 school year obtained through state department of public health (Table S1).

**Table S1.**
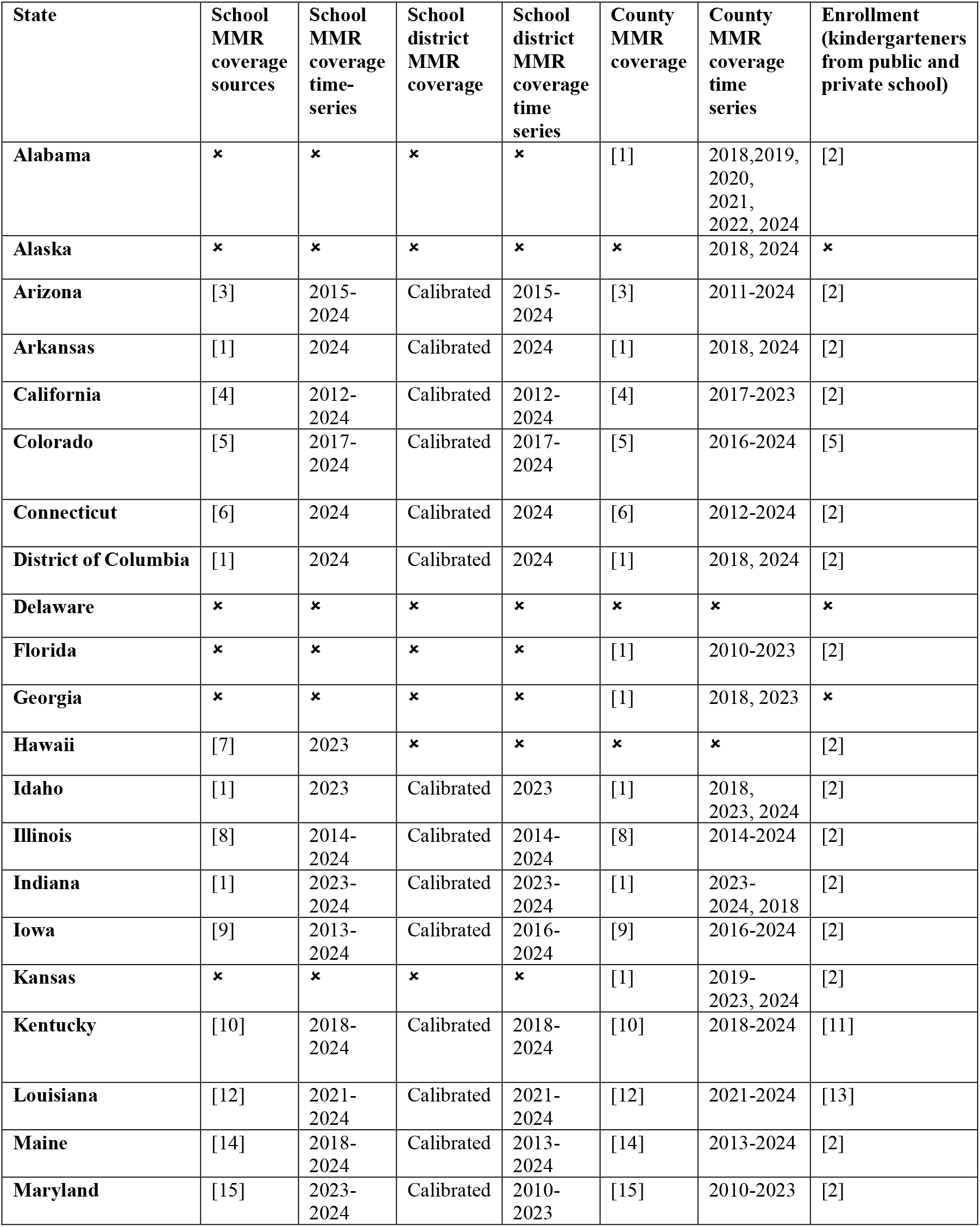
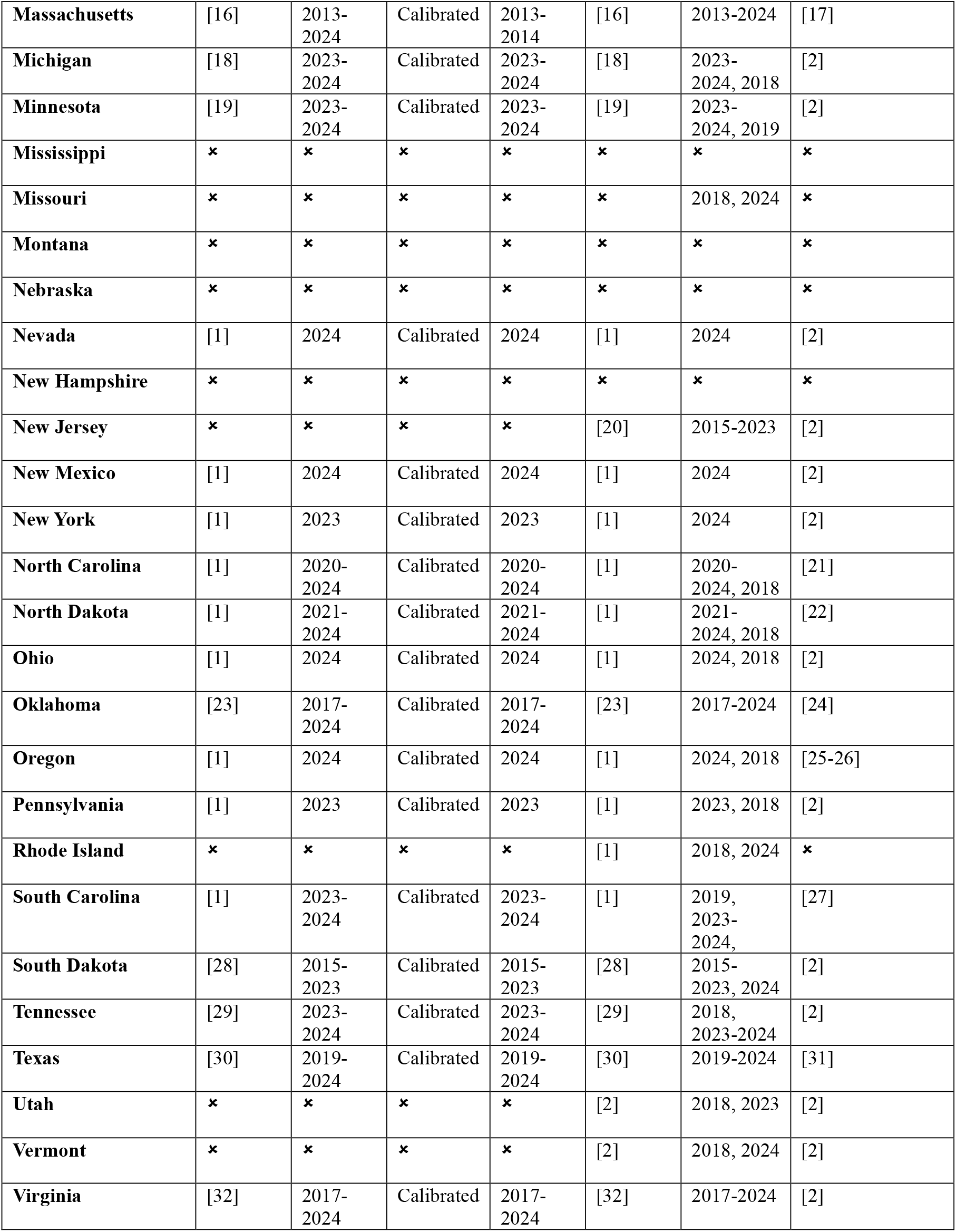
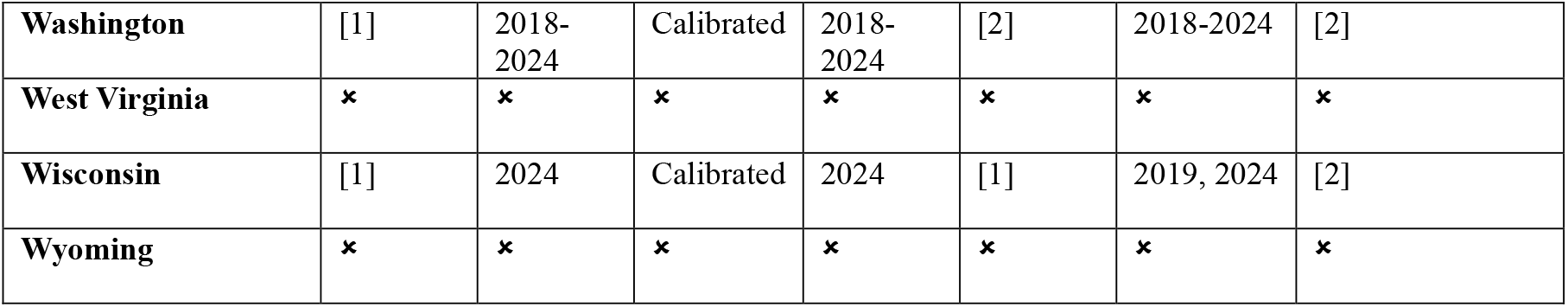
MMR vaccination coverage and enrollment data sources by state.

| State | School MMR coverage sources | School MMR coverage time-series | School district MMR coverage | School district MMR coverage time series | County MMR coverage | County MMR coverage time series | Enrollment (kindergarteners from public and private school) |
| --- | --- | --- | --- | --- | --- | --- | --- |
| Alabama | ✕ | ✕ | ✕ | ✕ | [1] | 2018,2019, 2020, 2021, 2022, 2024 | [2] |
| Alaska | ✕ | ✕ | ✕ | ✕ | ✕ | 2018, 2024 | ✕ |
| Arizona | [3] | 2015-2024 | Calibrated | 2015-2024 | [3] | 2011-2024 | [2] |
| Arkansas | [1] | 2024 | Calibrated | 2024 | [1] | 2018, 2024 | [2] |
| California | [4] | 2012-2024 | Calibrated | 2012-2024 | [4] | 2017-2023 | [2] |
| Colorado | [5] | 2017-2024 | Calibrated | 2017-2024 | [5] | 2016-2024 | [5] |
| Connecticut | [6] | 2024 | Calibrated | 2024 | [6] | 2012-2024 | [2] |
| District of Columbia | [1] | 2024 | Calibrated | 2024 | [1] | 2018, 2024 | [2] |
| Delaware | ✕ | ✕ | ✕ | ✕ | ✕ | ✕ | ✕ |
| Florida | ✕ | ✕ | ✕ | ✕ | [1] | 2010-2023 | [2] |
| Georgia | ✕ | ✕ | ✕ | ✕ | [1] | 2018, 2023 | ✕ |
| Hawaii | [7] | 2023 | ✕ | ✕ | ✕ | ✕ | [2] |
| Idaho | [1] | 2023 | Calibrated | 2023 | [1] | 2018, 2023, 2024 | [2] |
| Illinois | [8] | 2014-2024 | Calibrated | 2014-2024 | [8] | 2014-2024 | [2] |
| Indiana | [1] | 2023-2024 | Calibrated | 2023-2024 | [1] | 2023-2024, 2018 | [2] |
| Iowa | [9] | 2013-2024 | Calibrated | 2016-2024 | [9] | 2016-2024 | [2] |
| Kansas | ✕ | ✕ | ✕ | ✕ | [1] | 2019-2023, 2024 | [2] |
| Kentucky | [10] | 2018-2024 | Calibrated | 2018-2024 | [10] | 2018-2024 | [11] |
| Louisiana | [12] | 2021-2024 | Calibrated | 2021-2024 | [12] | 2021-2024 | [13] |
| Maine | [14] | 2018-2024 | Calibrated | 2013-2024 | [14] | 2013-2024 | [2] |
| Maryland | [15] | 2023-2024 | Calibrated | 2010-2023 | [15] | 2010-2023 | [2] |
| **Massachusetts** | [16] | 2013-2024 | Calibrated | 2013-2014 | [16] | 2013-2024 | [17] |
| **Michigan** | [18] | 2023-2024 | Calibrated | 2023-2024 | [18] | 2023-2024, 2018 | [2] |
| **Minnesota** | [19] | 2023-2024 | Calibrated | 2023-2024 | [19] | 2023-2024, 2019 | [2] |
| **Mississippi** | ✖ | ✖ | ✖ | ✖ | ✖ | ✖ | ✖ |
| **Missouri** | ✖ | ✖ | ✖ | ✖ | ✖ | 2018, 2024 | ✖ |
| **Montana** | ✖ | ✖ | ✖ | ✖ | ✖ | ✖ | ✖ |
| **Nebraska** | ✖ | ✖ | ✖ | ✖ | ✖ | ✖ | ✖ |
| **Nevada** | [1] | 2024 | Calibrated | 2024 | [1] | 2024 | [2] |
| **New Hampshire** | ✖ | ✖ | ✖ | ✖ | ✖ | ✖ | ✖ |
| **New Jersey** | ✖ | ✖ | ✖ | ✖ | [20] | 2015-2023 | [2] |
| **New Mexico** | [1] | 2024 | Calibrated | 2024 | [1] | 2024 | [2] |
| **New York** | [1] | 2023 | Calibrated | 2023 | [1] | 2024 | [2] |
| **North Carolina** | [1] | 2020-2024 | Calibrated | 2020-2024 | [1] | 2020-2024, 2018 | [21] |
| **North Dakota** | [1] | 2021-2024 | Calibrated | 2021-2024 | [1] | 2021-2024, 2018 | [22] |
| **Ohio** | [1] | 2024 | Calibrated | 2024 | [1] | 2024, 2018 | [2] |
| **Oklahoma** | [23] | 2017-2024 | Calibrated | 2017-2024 | [23] | 2017-2024 | [24] |
| **Oregon** | [1] | 2024 | Calibrated | 2024 | [1] | 2024, 2018 | [25-26] |
| **Pennsylvania** | [1] | 2023 | Calibrated | 2023 | [1] | 2023, 2018 | [2] |
| **Rhode Island** | ✖ | ✖ | ✖ | ✖ | [1] | 2018, 2024 | ✖ |
| **South Carolina** | [1] | 2023-2024 | Calibrated | 2023-2024 | [1] | 2019, 2023-2024, | [27] |
| **South Dakota** | [28] | 2015-2023 | Calibrated | 2015-2023 | [28] | 2015-2023, 2024 | [2] |
| **Tennessee** | [29] | 2023-2024 | Calibrated | 2023-2024 | [29] | 2018, 2023-2024 | [2] |
| **Texas** | [30] | 2019-2024 | Calibrated | 2019-2024 | [30] | 2019-2024 | [31] |
| **Utah** | ✖ | ✖ | ✖ | ✖ | [2] | 2018, 2023 | [2] |
| **Vermont** | ✖ | ✖ | ✖ | ✖ | [2] | 2018, 2024 | [2] |
| **Virginia** | [32] | 2017-2024 | Calibrated | 2017-2024 | [32] | 2017-2024 | [2] |
| **Washington** | [1] | 2018-2024 | Calibrated | 2018-2024 | [2] | 2018-2024 | [2] |
| **West Virginia** | × | × | × | × | × | × | × |
| **Wisconsin** | [1] | 2024 | Calibrated | 2024 | [1] | 2019, 2024 | [2] |
| **Wyoming** | × | × | × | × | × | × | × |

## Notes

### Competing Interest Statement

The authors have declared no competing interest.

### Author Declarations

Source data were openly available before the initiation of the study. All data and materials used in the analyses can be accessed through the paper.

